# Evaluating and Mitigating Limitations of Large Language Models in Clinical Decision Making

**DOI:** 10.1101/2024.01.26.24301810

**Authors:** Paul Hager, Friederike Jungmann, Kunal Bhagat, Inga Hubrecht, Manuel Knauer, Jakob Vielhauer, Robbie Holland, Rickmer Braren, Marcus Makowski, Georgios Kaisis, Daniel Rueckert

**Affiliations:** Lab for AI in Medicine, TUM, Munich, Germany; Institute for Diagnostic and Interventional Radiology, Klinikum rechts der Isar, TUM, Munich, Germany; Department of Medicine, ChristianaCare Health System, Wilmington, DE, USA; Department of Medicine III, Klinikum rechts der Isar, TUM, Munich, Germany; Department of Medicine II, University Hospital of the LMU Munich, Munich, Germany; Helmholtz Munich; Imperial College London

## Abstract

Clinical decision making is one of the most impactful parts of a physician’s responsibilities and stands to benefit greatly from AI solutions and large language models (LLMs) in particular. However, while LLMs have achieved excellent performance on medical licensing exams, these tests fail to assess many skills that are necessary for deployment in a realistic clinical decision making environment, including gathering information, adhering to established guidelines, and integrating into clinical workflows. To understand how useful LLMs are in real-world settings, we must evaluate them *in the wild*, i.e. on real-world data under realistic conditions. Here we have created a curated dataset based on the MIMIC-IV database spanning 2400 real patient cases and four common abdominal pathologies as well as a framework to simulate a realistic clinical setting. We show that current state-of-the-art LLMs do not accurately diagnose patients across all pathologies (performing significantly worse than physicians on average), follow neither diagnostic nor treatment guidelines, and cannot interpret laboratory results, thus posing a serious risk to the health of patients. Furthermore, we move beyond diagnostic accuracy and demonstrate that they cannot be easily integrated into existing workflows because they often fail to follow instructions and are sensitive to both the quantity and order of information. Overall, our analysis reveals that LLMs are currently not ready for clinical deployment while providing a dataset and framework to guide future studies.

## 1 Main

Large language models (LLMs) have the potential to revolutionize our medical system[54]. They can already streamline report generation and summarization[61, 59, 8, 37], answer biomedical questions with[60, 5, 59] and without[50, 49, 38] images, and could soon effectively interpret multimodal data for precision medicine in the clinic[6]. Importantly, as humans primarily interact with the world through language, LLMs are poised to be the point of access to the multimodal medical AI solutions of the future[36]. Until now, however, the diagnostic capabilities of models have been tested in structurally simple medical contexts, such as canonical vignettes of hypothetical patients or clinical case challenges. In both scenarios, all the required diagnostic information is provided upfront and there is a single answer to be selected from a list of options. This type of question dominates the medical licensing exams that have been used to test LLMs such as the United States Medical Licensing Exam (USMLE)[27, 26], Applied Knowledge Test (AKT) of the Membership of the Royal College of General Practicioners (RCGP)[53], and All India Institute of Medical Sciences (AIIMS) & National Eligibility cum Entrance Test Postgraduate (NEETS PG) entrance exam[42]. LLMs have proven their ability to excel in such scenarios, scoring well above passing on medical licensing exams[31, 21, 49, 50, 38, 56, 39] and rivaling clinician performance on clinical case challenges [34, 28, 10, 18].

However, while these medical licensing exams and clinical case challenges are suitable for testing the general medical knowledge of the test-taker, they are far removed from the daily task of clinical decision making. For medical practitioners, clinical decision making is one of the most important and complex everyday responsibilities. It is a multi-step process that requires gathering and synthesizing data from diverse sources and continuously evaluating the facts to reach an evidence-based decision on patient diagnosis and treatment[7, 55]. To reach a precise diagnosis, physicians must gather the necessary information based on the availability of diagnostic resources and adhere to established guidelines. Furthermore, they must carefully consider the patient-specific symptoms to plan the optimal treatment. As this process is very labor-intensive, there exists great potential in harnessing AI, such as LLMs, to alleviate much of the workload, ultimately aiming to autonomously, efficiently, and safely reach a final diagnosis which can then be confirmed by physicians[48, 3]. Thus, to understand how useful LLMs would be in such a real-world setting, they must be evaluated on real-world data and under realistic conditions. However, the only analysis that tested an LLM throughout the clinical workflow, used curated lists of possible answers and examined only 36 hypothetical clinical vignettes[46]. Furthermore, any model that is used in such a high-stakes clinical context must not only be highly accurate, but also adhere to diagnostic and treatment guidelines, be robust, and follow instructions, all of which have not been tested in previous medical evaluations.

Here, we present a curated dataset based on the Medical Information Mart for Intensive Care (MIMIC-IV) database spanning 2400 real patient cases and four common abdominal pathologies (appendicitis, pancreatitis, cholecystitis, diverticulitis) as well as a comprehensive evaluation framework around our dataset to simulate a realistic clinical setting. We provide LLMs with a patient’s history of present illness and ask them to iteratively gather and synthesize additional information such as physical examinations, laboratory results, and imaging reports until they are confident enough to provide a diagnosis and treatment plan. Our new dataset, task, and analysis represent the first large-scale evaluation of LLMs on every-day clinical decision making tasks in a realistic, open-ended environment. Unlike previous works, we test the autonomous information gathering and open-ended diagnostic capabilities of models, representing an essential step towards evaluating their suitability as clinical decision makers.

To understand how useful LLMs would be in the clinic today, we compare the diagnostic accuracy of the models on our dataset with that of clinicians. Furthermore, we propose and evaluate a new range of characteristics beyond diagnostic accuracy, such as adherence to diagnostic and treatment guidelines, correct interpretation of laboratory test results, instruction following capabilities, and robustness to changes in instructions, information order, and information quantity. Finally, we show that summarizing progress and filtering laboratory results for only abnormal results addresses some of the current limitations of models. We make our evaluation framework and dataset freely and openly available to guide future studies considering the use of LLMs in clinical practice.

## 2 Results

### 2.1 Creating the MIMIC-CDM dataset and evaluation framework

Our curated dataset, MIMIC Clinical Decision Making (MIMIC-CDM), is created using the well-established MIMIC-IV database, which is managed by the Massachusetts Institute of Technology (MIT). MIMIC-IV contains de-identified records of patient measurements, diagnoses, procedures, treatments, and free-text clinical notes such as discharge summaries and radiologists reports from patients admitted to the Beth Israel Deaconess Medical Center in Boston, USA, from 2008 to 2019[22]. Figure 1a and Materials Section 6 list the steps involved in creating the MIMIC-CDM dataset and its makeup. Our dataset contains electronic health record data from 2400 unique patients presenting with acute abdominal pain to the emergency department and whose primary diagnosis was one of the following pathologies: appendicitis, cholecystitis, diverticulitis, or pancreatitis. We chose these target pathologies as they represent clinically important diagnoses of a common chief complaint, abdominal pain, which accounts for 10% of all emergency department visits [11, 15]. Furthermore, different treatment strategies, ranging from antibiotics to surgery, are necessary depending on the severity of the condition. Thus, a thorough understanding of the specifics of a patient’s case is required to recommend optimal treatment. Importantly, a good differentiation between the four pathologies can be achieved using standard diagnostic tests, all of which are present in MIMIC-CDM.

**Figure 1:**
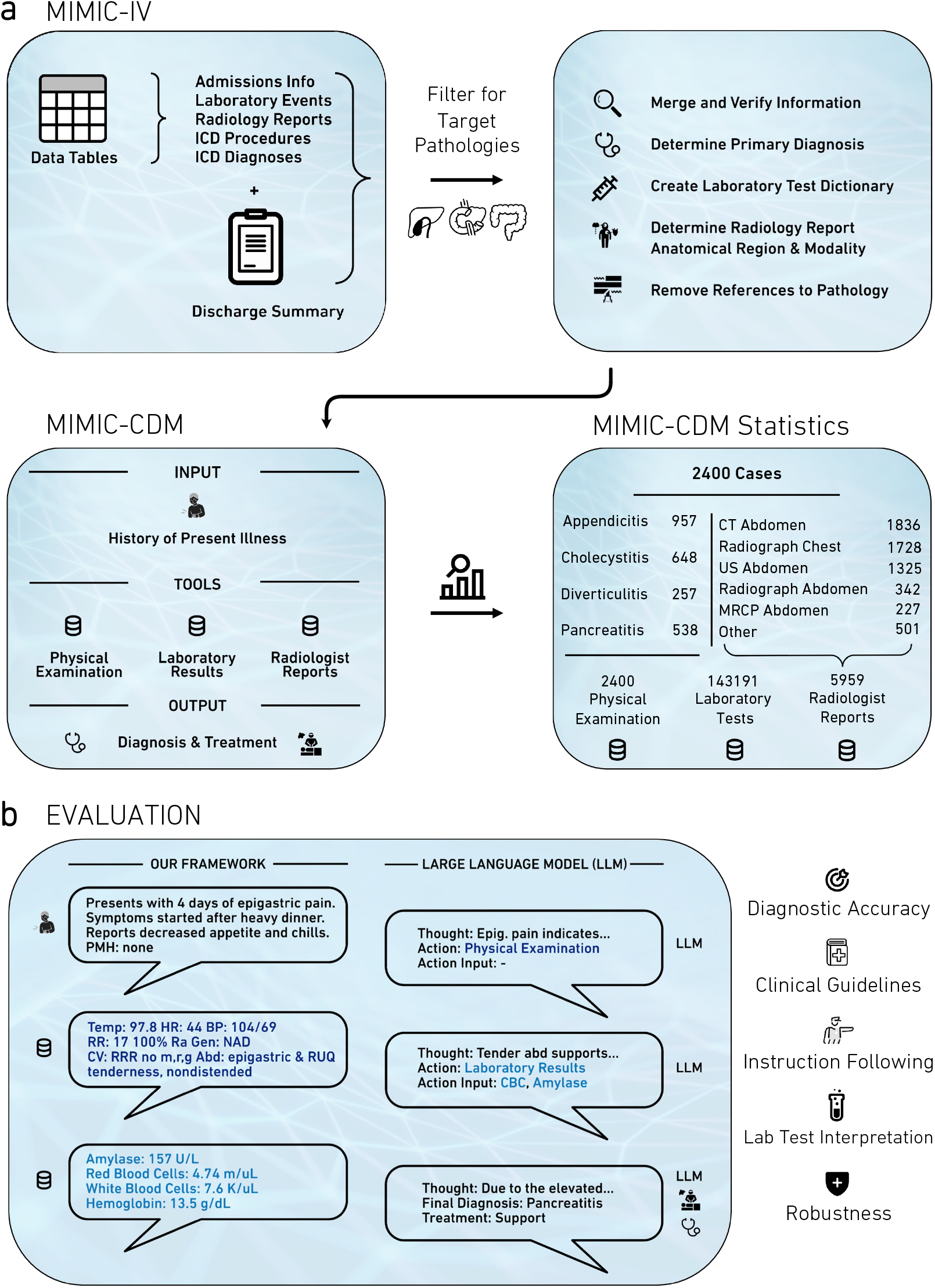
(a.) To properly evaluate LLMs for clinical decision making in realistic conditions, we created a curated dataset from real-world patient cases derived from the MIMIC-IV database, which contains comprehensive electronic health record data recorded during hospital admissions. (b.) Our evaluation framework reflects a realistic clinical setting and thoroughly evaluates LLMs across multiple criteria, including diagnostic accuracy, adherence to diagnostic and treatment guidelines, consistency in following instructions, ability to interpret laboratory results, and robustness to changes in instruction, information quantity, and information order. Abbreviations: ICD: International Classification of Diseases. CT: Computed Tomography. US: Ultrasound. MRCP: Magnetic resonance cholangiopancreatography.

To reflect a realistic clinical setting that allows LLMs to autonomously engage in every step of the clinical decision making process we have created a comprehensive evaluation framework around our dataset. Using our framework and dataset, we present LLMs with a patient’s history of present illness and task them to gather and synthesize information such as physical examinations, specific laboratory results, and distinct imaging reports to arrive at a diagnosis and treatment plan, as shown in Figure 1b. We then take the complete interaction and evaluate the LLM for diagnostic accuracy as well as adherence to diagnostic and treatment guidelines, and instruction following capabilities. For comparisons with practicing clinicians and further tests concerning robustness, we evaluate the diagnostic accuracy of LLMs when provided with all necessary information for a diagnosis; a dataset we call MIMIC Clinical Decision Making with Full Information (MIMIC-CDM-FI). In this dataset, we include the history of present illness, physical examination, relevant laboratory results, and all abdominal imaging, before directly asking for a diagnosis.

In our study, we tested the leading open-access LLM developed by Meta, Llama 2[58], and its derivatives. We focus on the largest model with 70 billion (B) parameters, as it performed best on benchmarks including medical questions. We test both generalist versions such as Llama 2 Chat (70B)[58], Open Assistant (OASST) (70B)[30], and WizardLM (70B)[63], as well as medical-domain aligned models such as Clinical Camel (70B)[56], and Meditron (70B)[12]. Further information on the models and our selection criteria can be found in Materials Section 6.3. Data taken from the MIMIC database is currently prohibited from being used with external APIs, such as that of OpenAI or Google, due to data privacy concerns and data usage agreements, so neither ChatGPT nor GPT-4 could be tested. Furthermore, multiple requests for access to Google’s MedPaLM models were denied. We note that Llama 2, Clinical Camel, and Meditron have been shown to match and even exceed ChatGPT performance on medical licensing exams and biomedical question answering tests[56, 12].

### 2.2 LLMs diagnose significantly worse than clinicians

To ensure patient safety, LLMs must diagnose at least as well as clinicians. Thus, we compared the diagnostic accuracy of the models on a subset of MIMIC-CDM-FI to four practicing hospitalists: two from the Klinikum Rechts der Isar Hospital of the Technical University of Munich, Germany (with two and three years of experience), one from the Ludwig Maximilian University Hospital in Munich, Germany, (four years of experience) and one from the Christiana Care hospital in Delaware, United States of America (29 years of experience). All four of the hospitalists are internal medicine physicians with emergency department experience. Each hospitalist was instructed to provide the primary pathology afflicting the patient and was given the same 100 patients in a random order to diagnose. 20 patients of each target pathology (appendicitis, cholecystitis, diverticulitis, pancreatitis) were included, with an additional five patients each of four other abdominal pathologies: gastritis, urinary tract infection, esophageal reflux, and hernia. Each LLM model was evaluated 10 times, using different random seeds, over the subset of 80 patients to increase statistical power. All statistical tests were corrected for multiple comparisons.

We find that current LLMs perform significantly worse than clinicians as each model’s mean performance, averaged over all four pathologies, is significantly lower than the mean of all clinicians (Doctors vs. Llama 2 Chat, p=8.06e-10; Doctors vs. OASST, p=1.10e-4; Doctors vs. WizardLM, p=6.18e-6; Doctors vs. ClinicalCamel, p=1.89e-4; Doctors vs Meditron, p=6.40e-5) (Fig. 2). The diagnostic accuracy of the clinicians also varied, with the German hospitalists in residency (Mean = 87.50% *±* 3.68%) performing slightly worse than the more senior US hospitalist (Mean = 92.50%), which can be attributed to differences in both experience and language, considering all text was in English. Most models were able to match clinician performance on the simplest diagnosis, appendicitis, where 3 out of 4 clinicians also correctly diagnosed 20 out of 20 patients. While the Meditron model matched or exceeded the other models at diagnosing appendicitis, diverticulitis, and pancreatitis patients, it failed for cholecystitis, diagnosing only one patient with ‘cholecystitis’ and the others primarily with ‘gallstones’ without specifying their location or inflammatory effects. This mirrors the general performance of the models, which may perform well on certain pathologies but currently lack the diagnostic range of human hospitalists. In a standard clinical scenario, where every diagnosis is a possibility, models must perform consistently across all pathologies of a single initial complaint, such as abdominal pain, to be useful.

**Figure 2:**
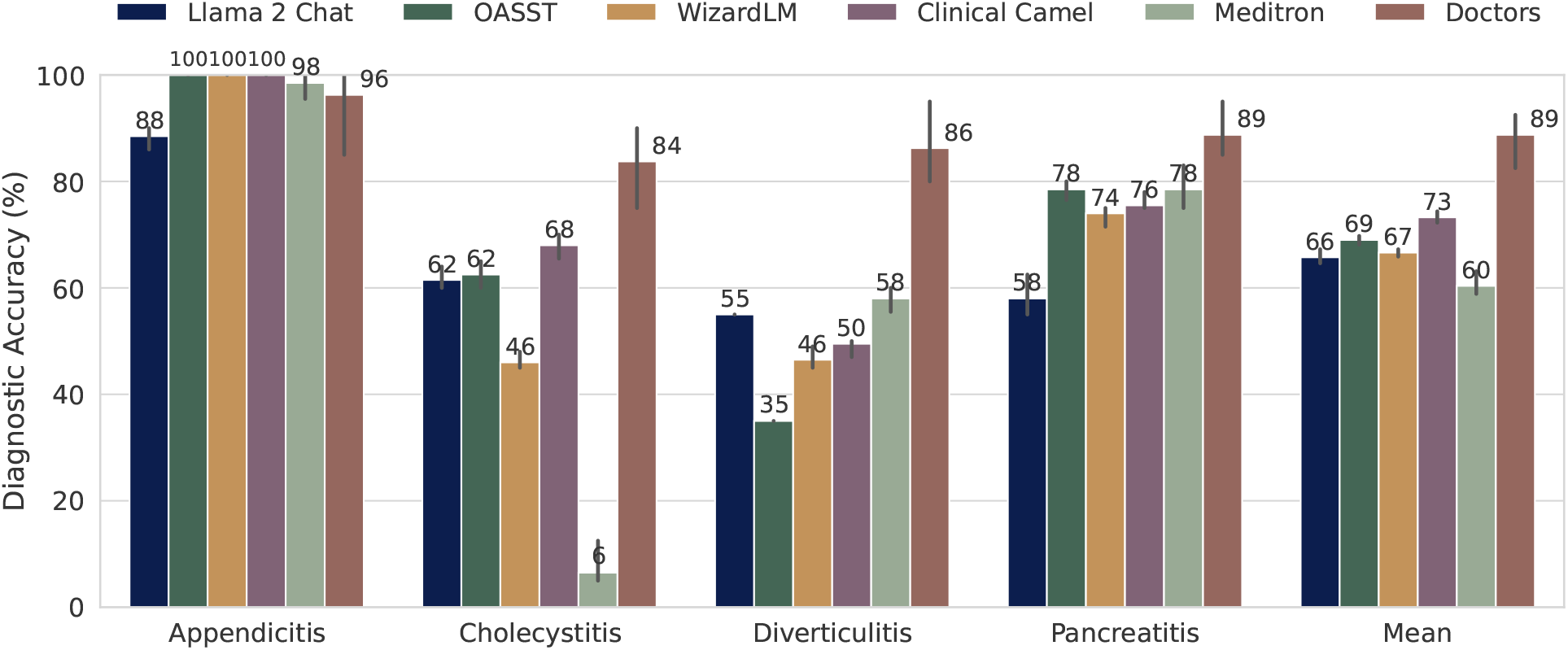
On a subset (n=80) of MIMIC-CDM-FI, LLMs perform significantly worse than doctors on average and especially on diseases such as cholecystitis and diverticulitis. The exact diagnostic accuracy is shown above each bar. Vertical lines indicate range of scores.

No specialist medical models performs significantly better on average than any generalist models (Clinical Camel vs. Llama 2 Chat, p=0.13; Clinical Camel vs. OASST, p=1.13, Clinical Camel vs. WizardLM, p=0.47; Meditron vs. Llama 2 Chat, p=1.11; Meditron vs. OASST, p=0.61; Meditron vs. WizardLM, p=1.04) (Fig. 2). Furthermore, as the medical LLMs are not instruction-fine-tuned (i.e. trained to understand and undertake new tasks), they are unable to complete the full clinical decision making task where they must first gather information and then come to a diagnosis. As this is the primary use-case of a clinical decision making model, we exclude them from all further analysis and only examine the Llama 2 Chat, OASST and WizardLM models for the rest of this work.

In our simulated clinical environment, which uses the MIMIC-CDM dataset, the LLM must specify all information it wishes to gather to accurately diagnose a patient. We observe a general decrease in performance, compared to MIMIC-CDM-FI (Supplementary Fig. 13), across the pathologies (Fig. 3). The mean diagnostic averages fall to 45.5% (vs. 58.8% on MIMIC-CDM-FI) for Llama 2 Chat, 54.9% (vs. 67.8%) for OASST, and 53.9% (vs. 65.1%) for WizardLM. Mirroring MIMIC-CDM-FI, all models performed best in diagnosing appendicitis (Llama 2 Chat: 74.6%, OASST: 82.0%, WizardLM: 78.4%) which is most likely due to the fact that appendicitis patients have consistent key symptoms with 791 of 957 radiologist reports (82.7%) directly stating that the appendix is dilated, enlarged, or fluid-filled, and typically lack other intra-abdominal pathology descriptions which distract from the acute diagnosis.

**Figure 3:**
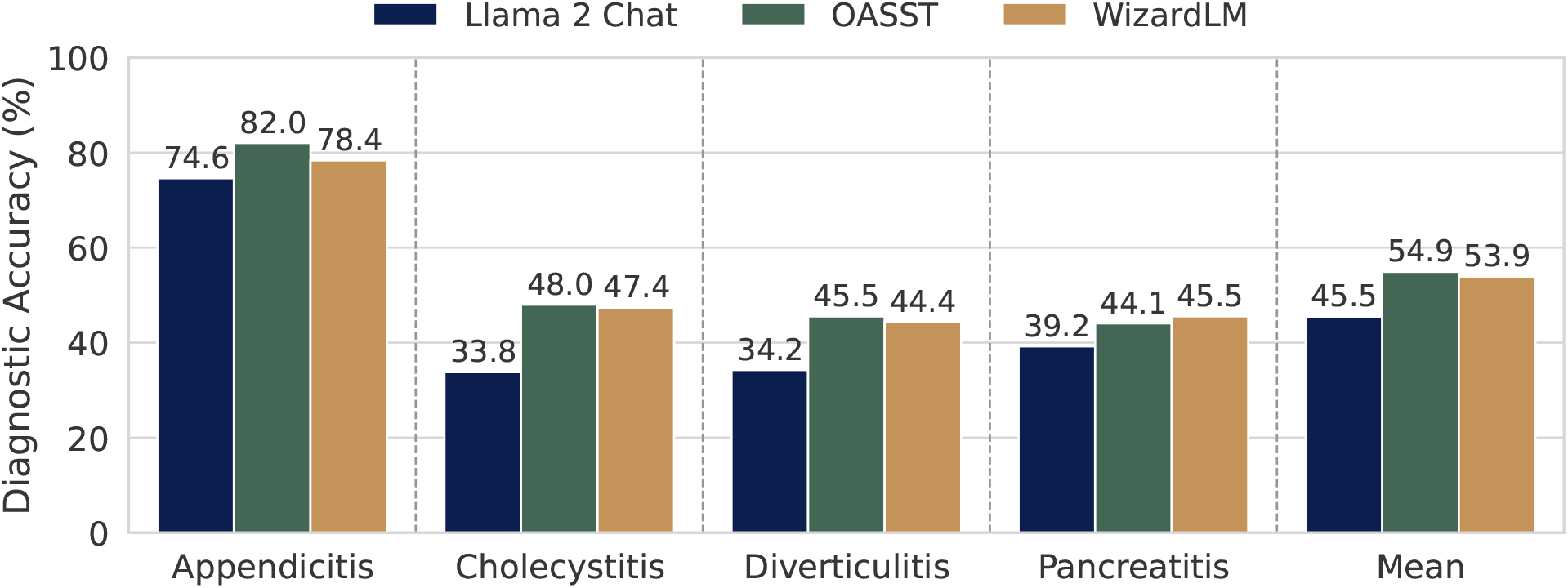
When tasked with gathering all information required for clinical decision making themselves, LLMs perform best when diagnosing appendicitis but perform poorly on the other three pathologies of cholecystitis, diverticulitis, and pancreatitis. In such a realistic clinical scenario, model performance decreased compared to the retrospective diagnosis with all information provided (MIMIC-CDM-FI). The exact diagnostic accuracy is shown above each bar.

In summary, LLMs do not reach the diagnostic accuracy of clinicians across all pathologies, especially when they must gather all information themselves. Thus, without extensive physician supervision, they would reduce the quality of care that patients receive and are currently unfit for the task of clinical decision making.

### 2.3 LLMs are hasty and unsafe clinical decision makers

In addition to poor diagnostic accuracy, LLMs often fail to order the exams required by diagnostic guidelines, do not follow treatment guidelines, and are incapable of interpreting lab results, making them a risk to patient safety.

To help clinicians consistently and safely reach a final diagnosis, diagnostic guidelines are published by associations of medical experts. The guidelines help guarantee a consistent standard of care by establishing a clinical consensus of which tests should be used to effectively diagnose a pathology, based on large clinical trials and clinician experience. The current clinical guidelines used for this study were: appendicitis[16], cholecystitis[44], diverticulitis [25], and pancreatitis[32].

All guidelines recommended physical examinations as an essential part of the diagnostic process, preferably as the first action. This is because they immediately provide valuable information as to the severity of the patient’s condition and a base for subsequent requests for laboratory tests and imaging. We find that only Llama 2 Chat consistently asks for physical examination results, either as the first action (97.1%) or at all (98.1%) (Supplementary Fig. 11). The other two models requested less examinations (OASST: 79.8% & 87.7%; WizardLM: 53.1% & 63.9%), thereby omitting an essential piece of information.

Hospitalists routinely order laboratory tests to measure biological markers and evaluate organ function, allowing them to track changes in a patient’s health and detect underlying biochemical shifts due to disease. Based on the diagnostic guidelines, we defined categories of necessary laboratory tests for each pathology, including signs of inflammation, functional fitness of the liver and gallbladder, pancreas enzymes, and the severity of a patient’s pancreatitis. The exact tests included in each category can be found in Supplementary Section D. We find that no model consistently orders all necessary tests, despite each test being independently requested by all doctors in the MIMIC-CDM dataset (Fig. 4). While OASST performs better than the other two models, reaching up to 93.3% and 87.2% in the inflammation category for appendicitis and diverticulitis, it often does not order the necessary tests for a diagnosis of pancreatitis (pancreas enzymes: 56.5%, severity: 76.2%), partially explaining why its diagnostic performance on pancreatitis was only 44.1% (Fig. 3). Not ordering all necessary tests makes it difficult to differentiate between abdominal pathologies, as laboratory results can provide an indication which organ is currently afflicted or functioning normally.

**Figure 4:**
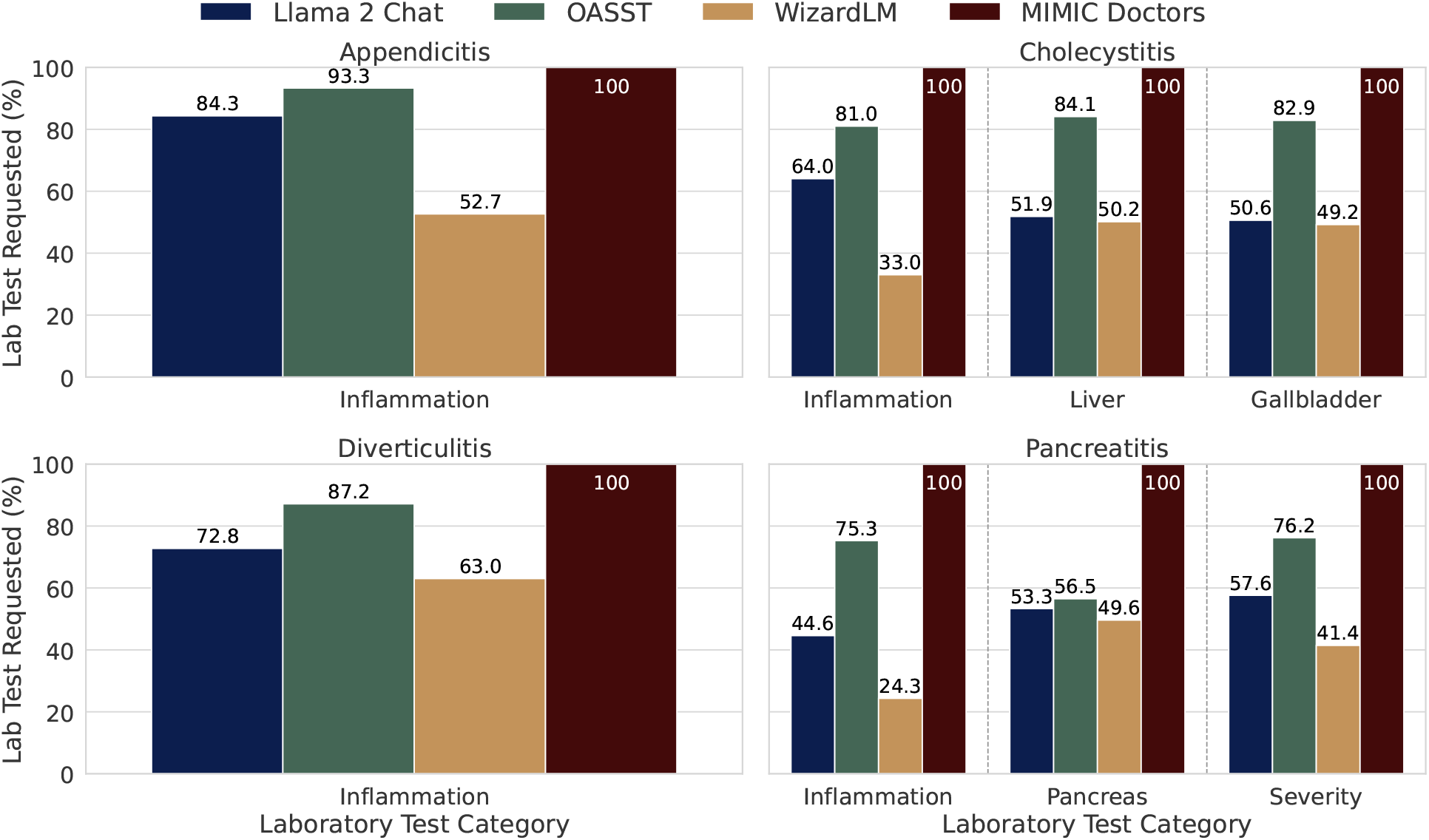
LLMs often do not order the necessary laboratory tests required to establish a diagnosis, despite each necessary test being ordered independently by all doctors in the MIMIC-CDM dataset. The tests, defined by current diagnostic guidelines, help differentiate abdominal pathologies, as results can indicate which organ is currently pathologically afflicted or functioning normally.

While it is important to order the correct laboratory tests, it is even more essential to correctly interpret them. The laboratory tests in MIMIC-IV have reference ranges included when applicable, so we tested the interpretation capabilities of the models by providing the test result with the accompanying reference range and asking them to classify each result as either below, within, or above the provided range. Any human with numerical literacy should be able to achieve perfect accuracy on such a task, however all LLMs perform very poorly, especially in the critical categories of low test results (Chat: 26.5%, OASST: 70.2%, WizardLM: 45.8%) and high test results (Chat: 50.1%, OASST: 77.2%, WizardLM: 24.1%) (Supplementary Fig. 15). Such a basic incomprehension of laboratory test results is a great risk to patient safety and must be resolved before LLMs become clinically useful.

While diagnostic guidelines also have recommendations for which imaging modality is best suited for establishing a diagnosis, the modality used in practice can vary based on current availability and the particularities of a patient’s case. We find that models sometimes match the modalities requested by the doctors in the dataset, but often come to a diagnosis without requesting an abdominal imaging scan (Supplementary Fig. 12). We include the first abdominal imaging modality recorded for each patient in MIMIC-CDM for comparison. As we later show that imaging is the most useful diagnostic tool for the LLMs for all pathologies except pancreatitis (Fig. 8), occasional failure to request imaging could be partly responsible for their low diagnostic accuracy.

Taken together, the lack of consistency of the LLMs in ordering all of the required tests for a diagnosis based on current guidelines indicates a tendency to diagnose before understanding or considering all of the facts of the patient’s case. This could pose a risk to patients’ health as they would be diagnosed based on insufficient information.

In addition to not following diagnostic guidelines, LLMs generally fail to adhere to treatment guidelines (Fig. 5). To evaluate their ability to recommend appropriate treatments, we used the aforementioned guidelines to extract the possible treatments for each pathology and then to classify each treatment as either essential (e.g. antibiotics, support) or case specific (e.g. appendectomy, cholecystectomy, drainage). For each patient, we then determined if the case specific treatment was appropriate by matching against the actual operations performed on that patient, read from the MIMIC-CDM dataset. We evaluate a model’s treatment recommendation only when it correctly diagnosed a patient since an inaccurate diagnosis likely leads to inappropriate treatment. Fig. 5 shows the total counts of each treatment in MIMIC-CDM under each treatment name and the counts of each models’ correct diagnoses under each bar. We find that the LLMs consistently do not recommend appropriate and sufficient treatment, especially for patients with more severe forms of the pathologies. While they are consistent in recommending some treatments such as appendectomy for appendicitis (Chat: 97.5%, OASST: 97.5%, WizardLM: 96.9%) and antibiotics for diverticulitis (Chat: 88.6%, OASST: 97.4%, WizardLM: 86.0%), they rarely recommend other treatments when appropriate such as colectomy for diverticulitis patients with perforated colons (Chat: 22.2%, OASST: 0.0%, WizardLM: 10.0%) or drainage of infected pancreatic necrosis (Chat: 0.0%, OASST: 0.0%, WizardLM: 0.0%) and abscesses near the diverticula (Chat: 16.7%, OASST: 22.2%, WizardLM: 0.0%). Furthermore, they drastically under-treat appendicitis with regards to the necessity of antibiotics (Chat: 8.5%, OASST: 26.0%, WizardLM: 6.0%) and providing support (Chat: 4.3%, OASST: 9.9%, WizardLM: 2.7%), diverticulitis with the need for a colonoscopy in the future to check for colon cancer (Chat: 4.5%, OASST: 18.8%, WizardLM: 9.6%), and pancreatitis with sufficient support (where we expect mentions of fluids, pain management, and monitoring) (Chat: 39.8%, OASST: 71.3%, WizardLM: 42.9%). In summary, following the treatment recommendations of the models would negatively impact the health of patients, particularly those with more advanced stages of disease where indications for emergency operations were ignored.

**Figure 5:**
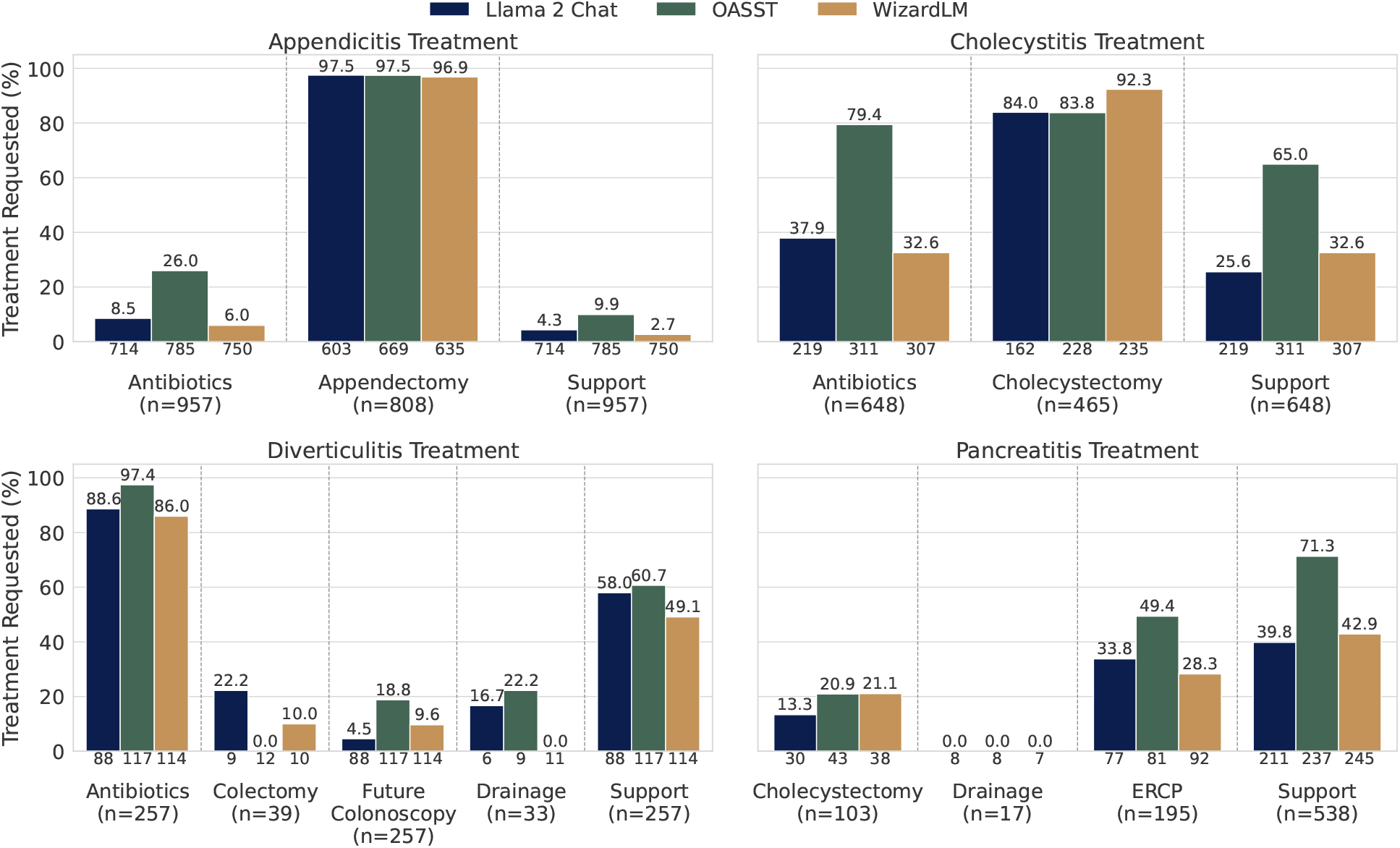
We find that LLMs do not consistently recommend essential and patient-specific treatment, especially for patients with more severe forms of the pathologies. We only score models on the subset of patients that they correctly diagnosed and that actually received a specific treatment. For example, of the 957 appendicitis patients, 808 received an appendectomy (indicated below the treatment name). Of those 808 patients, Llama 2 Chat correctly diagnosed 603 (indicated below the Llama 2 Chat bar). Of those 603 patients, Llama 2 Chat correctly recommended an appendectomy 97.5% of the time.

The tendency of LLMs to not gather the information required by diagnostic guidelines before returning a final diagnosis and treatment plan is particularly problematic considering their low overall diagnostic and treatment accuracy. Such hasty decision making combined with their poor diagnoses and treatment recommendations pose a serious risk to the health of patients without extensive clinician supervision and control.

### 2.4 LLMs require extensive clinician supervision to effectively integrate into clinical workflows

In addition to consistently and safely arriving at the correct diagnosis and treatment plan, models must integrate into established clinical workflows to be useful. Central to this is the ability to follow instructions and generate answers so they can be easily processed and used by other parts of the clinic without physician supervision. During the clinical decision making process, we provide clear instructions to the models as to how they should provide their requests and diagnosis, as well as which tools are available to them (see Materials Section 6.2). For example, diagnostic tools must be written in the ‘Action:’ field and desired tests must be specified in the ‘Action Input:’ field, and not in the middle of a paragraph surrounded by other text. This is essential to ensure that the desired tests can be consistently extracted so no manual clinician supervision and interpretation is required. Through extensive comparisons of LLM outputs with dictionaries of known exams and their synonyms, we go to great lengths to understand what tests are requested, even if the models do not follow our schema, recording every time they fail to follow instructions. We investigate the capabilities of models to follow our instructions at three timepoints during our analysis: when providing the next action to take, when requesting a tool, and when providing a diagnosis.

All models struggle to follow the provided instructions (Fig. 6), making errors every two to four patients when providing actions (Chat: 1.91, OASST: 3.90, WizardLM: 2.13) and hallucinating non-existent tools every two (Chat: 2.20 and WizardLM: 2.41) to five patients (OASST: 5.48). When providing diagnoses on the MIMIC-CDM dataset, errors are made every three to five patients (Chat: 3.18, OASST: 4.55, WizardLM: 3.64) while on the MIMIC-CDM-FI dataset there is a greater discrepancy in the number of patients diagnosed without formatting error (Chat: 1.12, OASST: 6.60, WizardLM: 28.21). While many of these errors are easily caught (Supplementary Table 6), the error rate is so high that extensive manual controls would be necessary to ensure model output is being correctly interpreted. Such poor abilities to follow instructions greatly reduces their usefulness in clinical systems as they would require large amounts of manual supervision to ensure proper performance.

**Figure 6:**
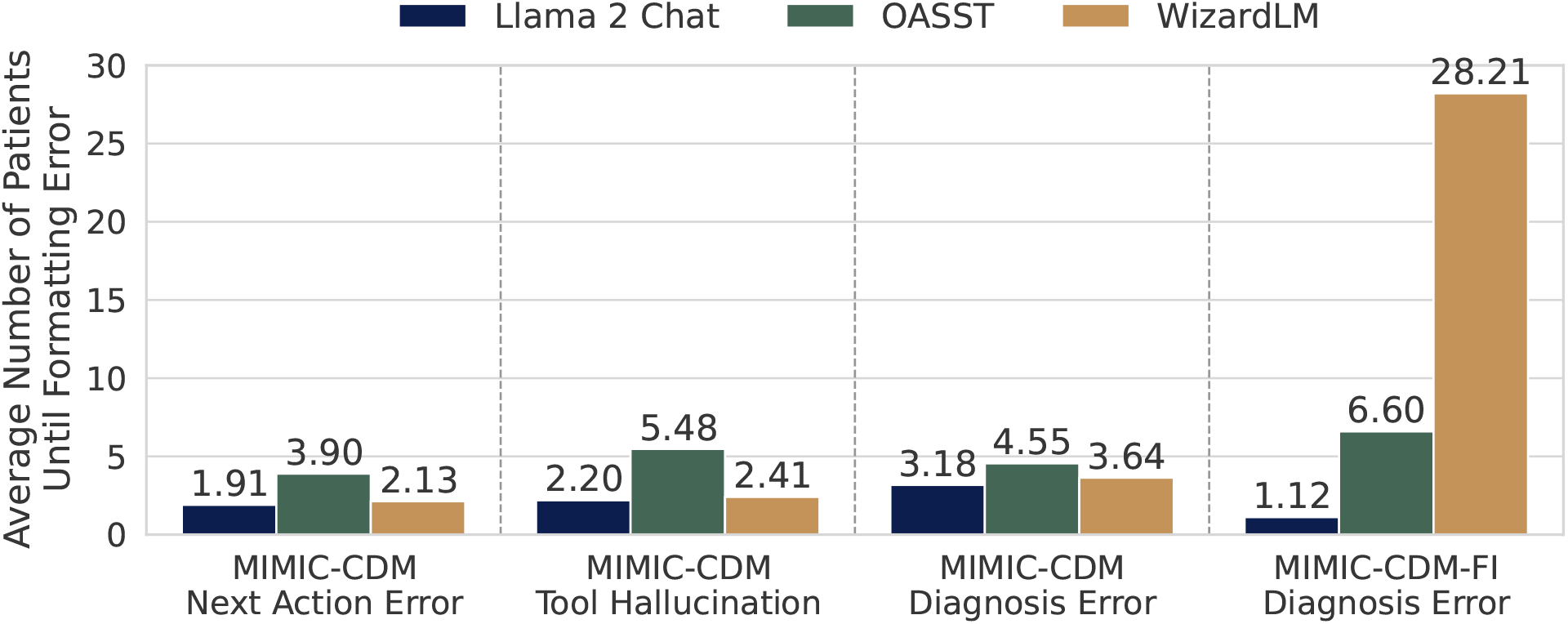
LLMs struggle to follow instructions, often introducing errors when providing the next action to take and hallucinating non-existent tools, up to once every two patients. Formatting errors while providing the diagnosis also regularly occur. In the clinic, extensive manual supervision would be required to ensure proper performance.

Another key component that must be fulfilled before we consider integrating such models into real world workflows is robustness. Models must not be sensitive to small changes in user instructions as their performance will then vary greatly based on who is interacting with them. On the MIMIC-CDM-FI dataset, we find that changes in instructions (Supplementary B) can lead to large changes (both positive and negative) in diagnostic accuracy (Fig. 7). For example, large changes are seen when removing system and user instructions (up to +5.1% for Chat on cholecystitis, down to –16.0% for Chat on pancreatitis), or when removing all medical terminology from the system instruction (up to +6.2 for WizardLM on diverticulitis, down to –3.5% for OASST on pancreatitis). Additionally, we see that even minor changes in instructions can greatly change diagnostic accuracy such as asking for the ‘Main Diagnosis:’ (up to +7.0% for Chat on diverticulitis, down to –10.6% for WizardLM on cholecystitis) or ‘Primary Diagnosis:’ (up +8.7% for Chat on Pancreatitis, down to –5.2% for WizardLM on cholecystitis) instead of ‘Final Diagnosis:’. Models should be able to provide the most appropriate diagnosis given the situation, in this case the reason for the patient’s abdominal pain, and not be sensitive to minute changes in phrasing so as not to require extensive clinician training before use.

**Figure 7:**
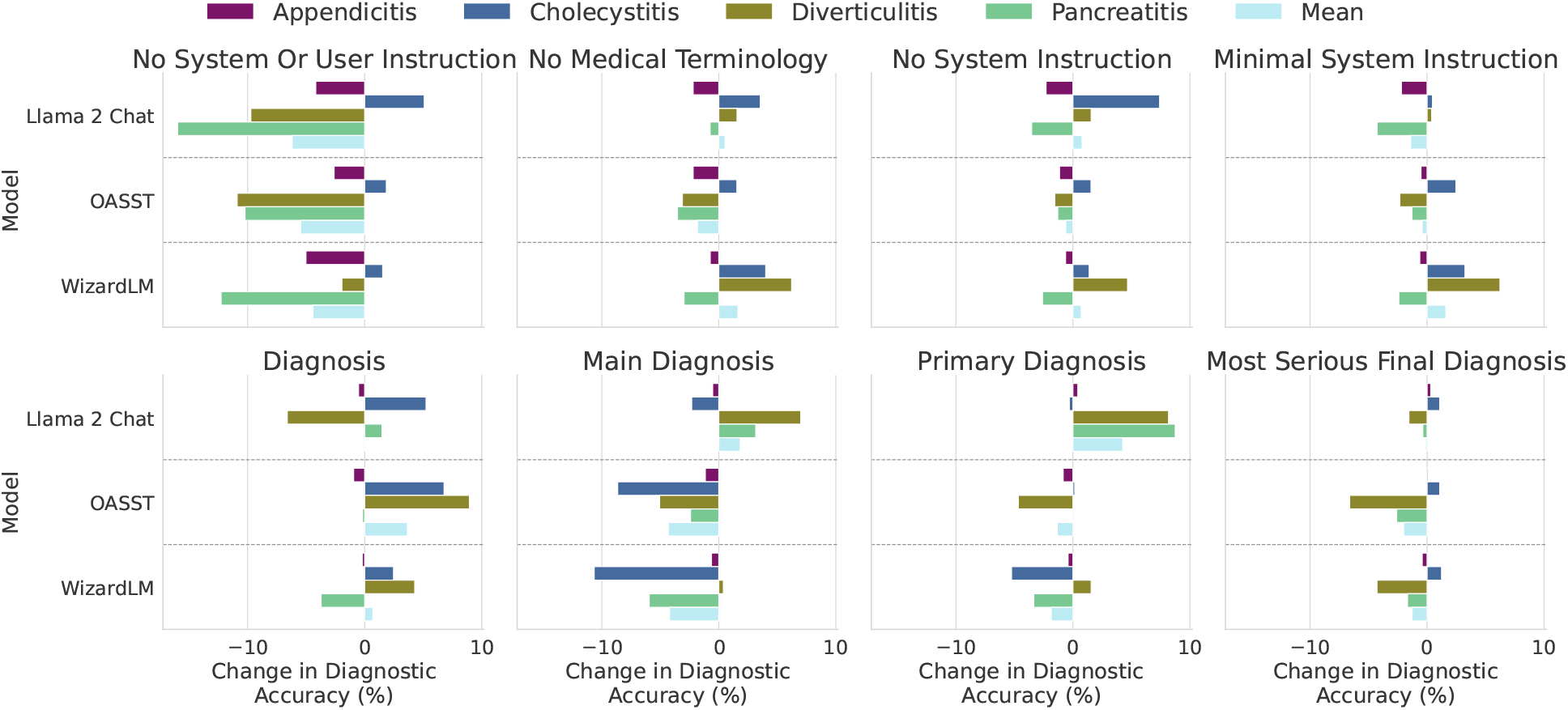
Often small changes in instructions, such as changing final diagnosis to main diagnosis or primary diagnosis, greatly affects the performance of the LLMs on the MIMIC-CDM-FI dataset. This would vary the quality of responses received depending on who is using the model.

Furthermore, LLMs must perform better or equally well when provided with more information. Large amounts of data are gathered during a patient’s hospital stay, with the patients in our dataset averaging 150 laboratory tests and 3 imaging exams per admission. Models must be able to focus on the key facts of the case to make a diagnosis. We show that models perform worse when all diagnostic exams are provided, typically attaining their best performance when only a single exam is provided in addition to the history of present illness (Fig 8). Removing information greatly increases diagnostic accuracy, with cholecystitis diagnosis improving by 18.5% for the Chat and 16.5% for the WizardLM models when only providing radiologist reports, and pancreatitis diagnosis improving by 21.6% (Chat), 9.5% (OASST), and 8.6% (WizardLM) when only providing laboratory results. This greatly reduces the usefulness of such models as they cannot simply be given all relevant information and be trusted to arrive at their best diagnosis. To optimize model performance, clinicians would have to decide which diagnosis is most likely to effectively filter the information presented. While such filtering by clinicians would increase their mean scores to a theoretical best of Chat: 72.2%, OASST: 70.9%, WizardLM: 71.8%, it would remove any benefit of deploying such a clinical decision making model.

**Figure 8:**
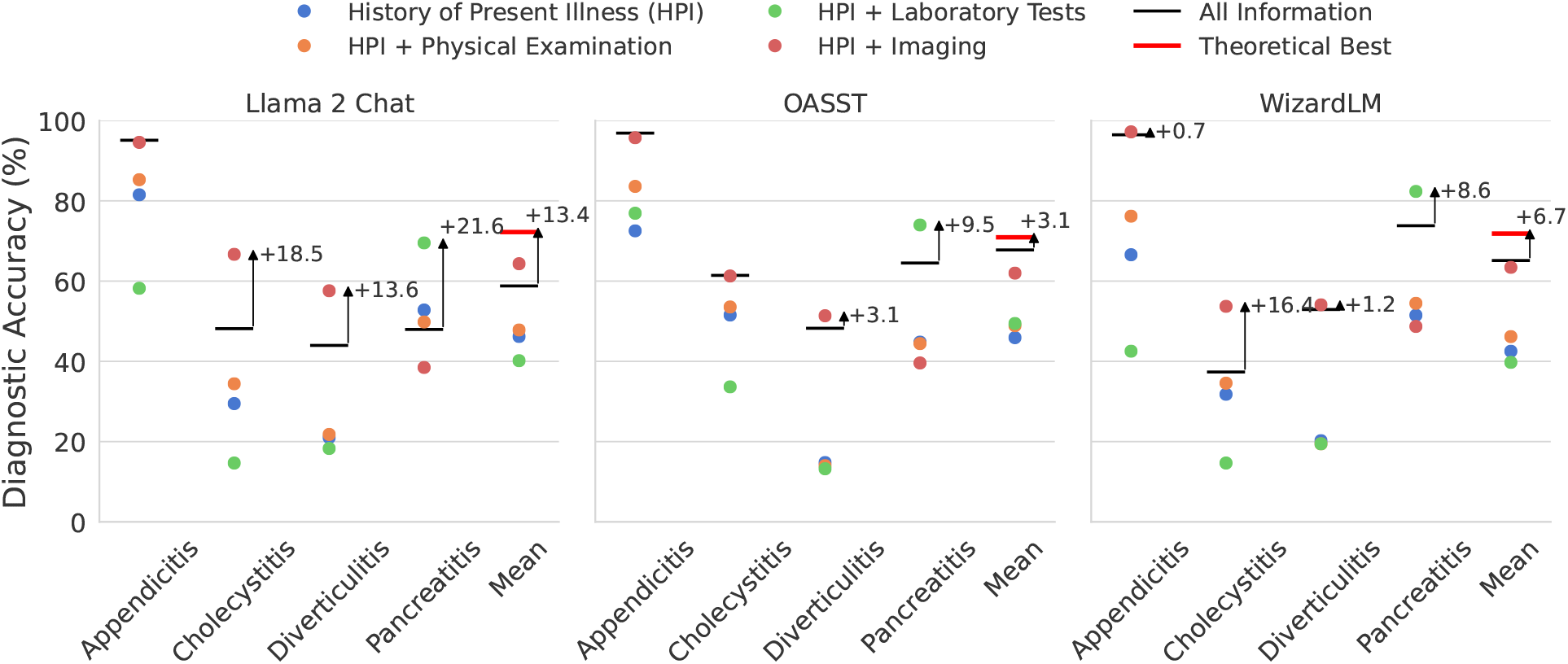
For almost all diseases, providing all information does not lead to the best performance on the MIMIC-CDM-FI dataset. This suggests that LLMs cannot focus on the key facts and degrade in performance when too much information is provided. This is a problem in the clinic where an abundance of information is typically gathered to wholistically understand the patients health and being able to focus on key facts is an essential skill. The theoretical best shows the mean performance if a clinician were to select the best diagnostic test for each pathology.

Models must also provide the same diagnosis, irrespective of the order in which the information is presented. We test the diagnostic consistency of the models on the MIMIC-CDM-FI dataset by switching the order of the information from the canonical physical examination, then laboratory tests, then imaging, to all possible permutations thereof (history of present illness is always included first). We show that all models have large ranges of performance, up to 18.0% (Chat – Pancreatitis), 7.9% (OASST – Cholecystitis), and 5.4% (WizardLM – Cholecystitis) (Fig. 9 and Supplementary Fig. 16). Importantly, we find that the order of information that delivers the best performance for each model is different for each pathology (Supplementary Section I). This again reduces the benefits of deploying the models as clinicians must constantly consider and monitor in which order they provide the models with information, in a disease specific manner, to not degrade performance.

**Figure 9:**
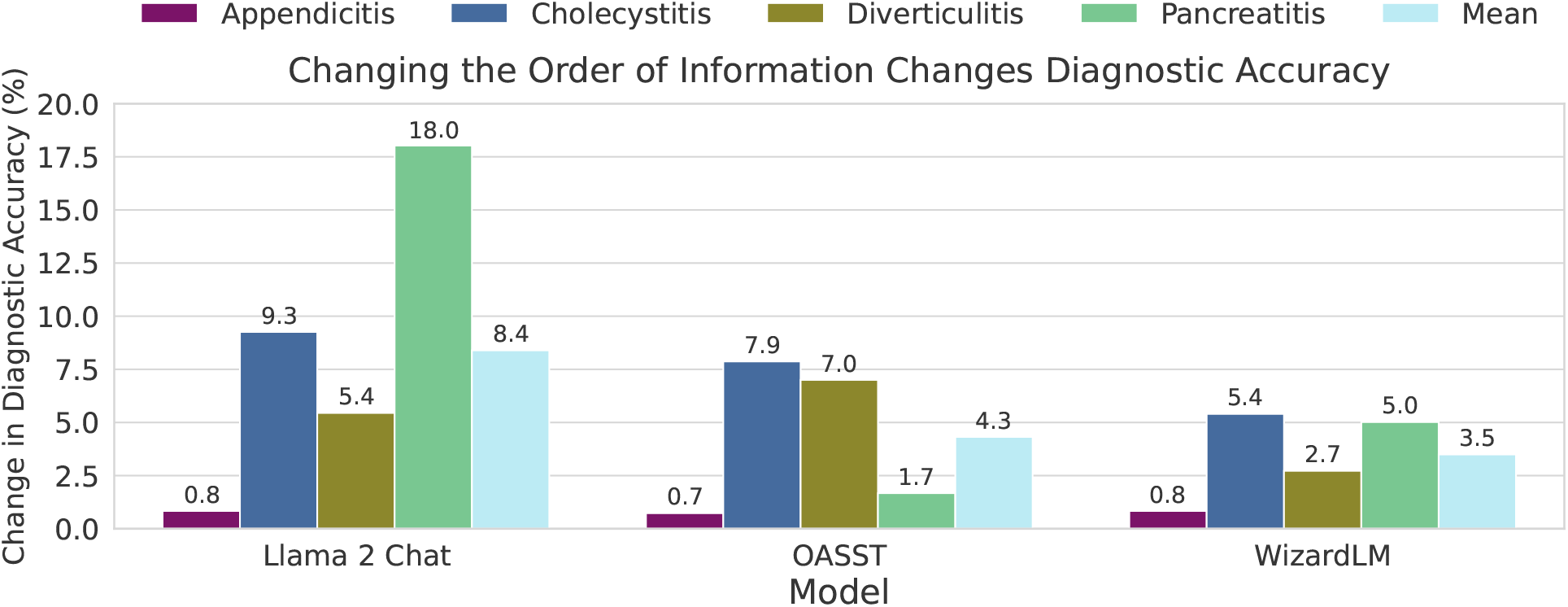
By mixing the order in which information is presented to LLMs, their performance changes despite the information included staying the same. This places an unnecessary burden upon clinicians who would need to make preliminary diagnoses to decide the order in which they feed the models with information for best performance.

In summary, extensive clinician supervision and prior evaluation of the most probable diagnosis would be required to ensure proper functioning of LLMs because they do not reliably follow instructions, perform better with a disease dependant order of information and degrade in performance when given relevant information. Furthermore, their sensitivity to small changes in instructions that seem inconsequential to humans would require extensive clinician training to ensure good performance.

### 2.5 Summarization and filtering for abnormal laboratory results partially mitigates limitations of current LLMs

To help address some of the limitations found in this analysis, we explore simple modifications that can be done without retraining the model. One major limitation is that LLMs are currently limited in the amount of text they can read, with all models tested in this study having a limit of 4096 tokens or approximately 2400 words. Due to this, if the history of present illness or radiologist reports are disproportionately long, the limit of the model is sometimes reached. To alleviate this, we implement an automatic summarization protocol that monitors the length of the text that the model receives. Once it approaches its maximum token amount, we ask the model to summarize the test results it has received, combining the healthy observations and emphasizing the pathological indications (Supplementary B.2). Removing such a summarization protocol resulted in marginal but consistent losses on the mean of –1.3% (Chat), –0.8% (OASST), and –0.5% (WizardLM), and particularly hurt the diagnosis of diverticulitis –4.7% (Chat), –2.7% (OASST), –3.5% (Wizard) (Supplementary Fig. 17).

Due to the inability of LLMs to reliably interpret laboratory results (Supplementary Fig. 15), even when provided with reference ranges, and their issues understanding larger quantities of information (Fig. 8), we find that filtering the laboratory results and removing all normal test results generally improved performance on the MIMIC-CDM-FI Dataset (Fig. 10). This increases the amount of other information that can be included before reaching the input length limit of the LLMs and reduces the amount of healthy information, which tends to confuse the tested models. Removing this information increased Llama 2 Chat and WizardLM mean performance by 6.0% and 3.9% respectively, while slightly changing mean OASST diagnostic accuracy by –0.9%. On the MIMIC-CDM-FI dataset, cholecystitis patients benefited the most from such a filter, with all models increasing in diagnostic accuracy (Chat: +16.8%, OASST: +3.5%, WizardLM: +14.8%). Pancreatitis patients in MIMIC-CDM-FI on the other-hand saw a light decrease in performance for the OASST (–6.1%) and WizardLM (–4.1%) models, with a slight increase for Llama 2 Chat (+2.0%). As many of our other analyses examine the general behaviour of laboratory tests and their impact on model performance, we do not use this fix for any other sections of this work. While this filtering improves the performance of the LLMs as they function today, ideally a model would perform best with all available information. Healthy laboratory test results are an important source of information for clinicians and should not degrade model performance.

**Figure 10:**
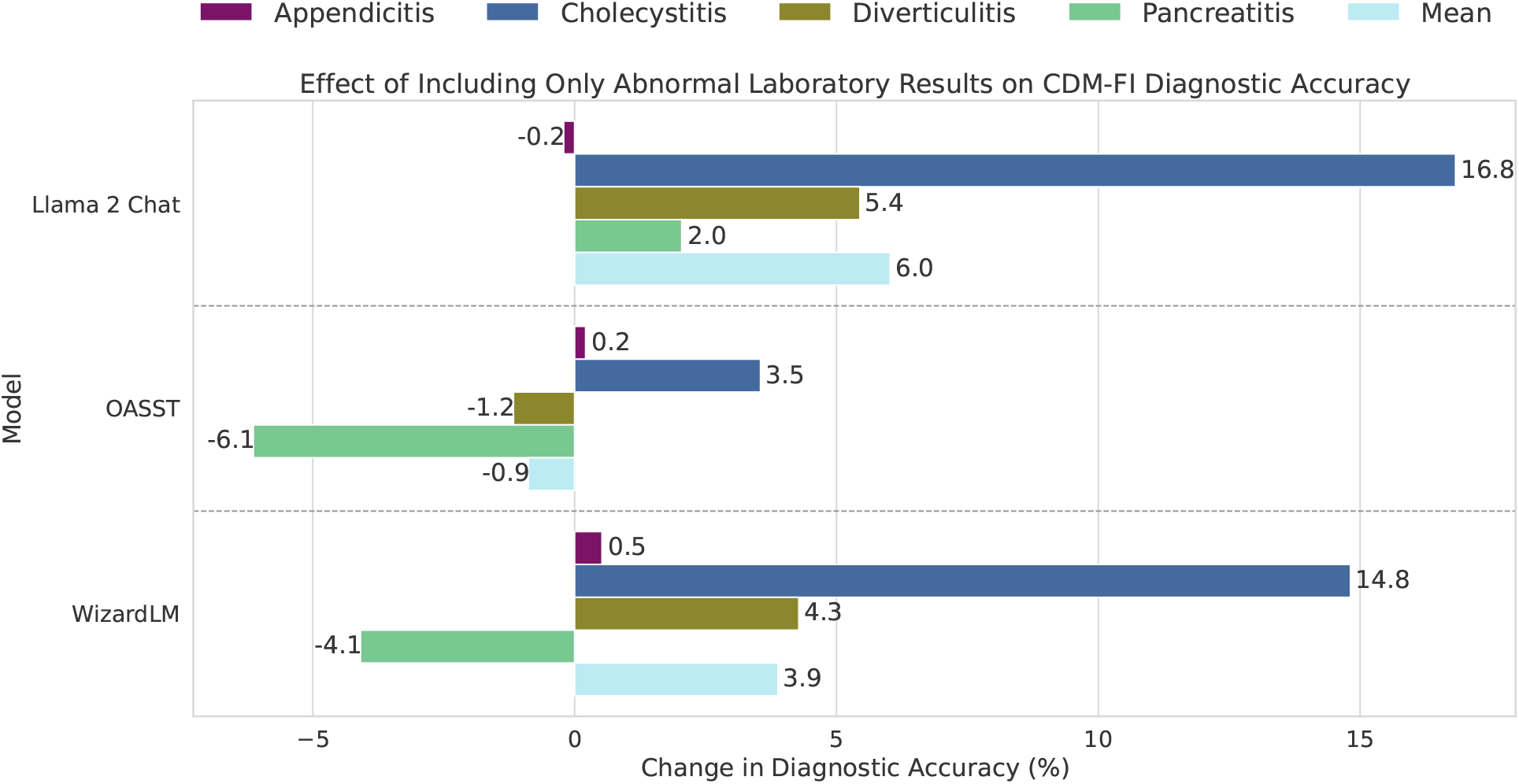
Filtering to include only abnormal laboratory results using the laboratory reference ranges provided in MIMIC-IV database generally improves model performance, especially for the cholecystitis pathology. This allows the model to focus more on abnormal, pathological signals.

## 3 Discussion

The strong performance of LLMs on medical licensing exams has led to increased interest in using them in clinical decision making scenarios involving real patients. However, medical licensing exams do not test the capabilities required for real world clinical decision making. We have, for the first time, evaluated leading open access large language models on making open-ended clinical decisions with thousands of real world patient cases to assess their potential benefits and possible harms. By not only comparing their diagnostic performance against clinicians, but also testing their information gathering abilities, adherence to guidelines, and instruction following capabilities as well as their robustness to changes in prompts, information order, and information quantity, we move for the first time beyond simple evaluations of diagnostic accuracy and establish a range of characteristics that are necessary for safe and robust integration into the clinic. In this work, we have shown that current leading LLMs are unsuitable for clinical decision making on all of these accounts.

Among the models tested in this study, we find that OASST performed best overall as it had decent diagnostic accuracy, generally requested appropriate laboratory exams, and was most robust to changes in information quantity. Llama 2 Chat had the worst overall diagnostic accuracy, often refused to follow instructions, and was heavily influenced by the order and quantity of information, but was the only model to consistently ask for a physical examination. WizardLM was the most robust to changes in the order of diagnostic exams and followed instructions well when returning diagnoses, but was the worst at following diagnostic guidelines, failing to consistently order physical examinations and necessary laboratory tests. Despite the performance of OASST being generally better than Chat and WizardLM across the diverse set of analyses included in this study, it is still not currently suitable for clinical use due to its inferior performance compared to clinicians, broad failure to order correct treatments, and general lack of robustness. While one of the medical domain models, Clinical Camel, achieved the highest diagnostic accuracy (mean=73% vs OASST mean=68%, Supplementary Fig. 13), its inability to participate in the iterative clinical decision making task precluded it from evaluations of its robustness and consistency which we believe to be essential to ensure safe deployment in the clinic. Other LLMs such as ChatGPT, GPT-4, Med-PALM, and Med-PALM 2 could not be tested due to the data privacy and usage agreements of MIMIC-IV, highlighting the risk of using corporate models in a sensitive area such as medicine, where patient privacy, transparency, and reliability are essential[57, 24].

The biggest barrier to using current LLMs in clinical practice is that no model consistently reached the diagnostic accuracy of clinicians across all pathologies, with a further decrease in accuracy when having to gathering diagnostic information themselves. A major weakness of these models is that their diagnostic accuracy is disease dependent, with markedly higher accuracy on appendicitis compared to cholecystitis, diverticulitis, and pancreatitis. A clinical model must diagnose well on all major differential diagnoses of a chief complaint, such as abdominal pain, to be useful. Additionally, LLMs are unable to classify a lab result as normal or abnormal, even when provided with its reference range. This is underscored by the fact that presenting the model with only abnormal laboratory results generally improved diagnostic performance.

We further found that the models do not follow diagnostic guidelines, which is particularly problematic considering their low overall diagnostic accuracy, indicating a tendency to diagnose before fully understanding a patient’s case. Insufficient diagnostic information also negatively affected the treatment recommendations of LLMs, where we showed that models do not follow all established treatment guidelines, especially for severe cases. The hasty decision making of the models combined with their low diagnostic performance and poor treatment recommendations pose a serious risk to the health of patients without extensive clinician supervision and control.

Beyond diagnostic accuracy, we extensively test models on their reliability and robustness which are essential characteristics to ensure consistent and safe patient care. We found that models struggle to follow instructions, often hallucinating non-existent tools and requiring continuous manual supervision to ensure proper performance. Models are also sensitive to seemingly inconsequential changes in instruction phrasing, requiring clinicians to carefully monitor the language they use to interact with the models to not degrade performance. Contrary to expectation, LLMs diagnose best when only a single diagnostic exam is provided rather than when given all relevant diagnostic information, demonstrating an inability to extract the most important diagnostic signal from the evidence. Counterintuitively, we found models to be sensitive to the order in which information is presented, resulting in large changes in diagnostic accuracy despite identical diagnostic information. Importantly, all of these weaknesses are disease-specific within each model, meaning that a different instruction, diagnostic test, and order of tests achieved best results for each pathology. As physicians would have to perform preliminary diagnostic evaluations in an attempt to maximize model performance according to their suspected diagnosis, all benefits of a clinical decision making system would be lost.

Many of the current limitations of LLMs exposed in our study have been shown concurrently in domains outside of medicine. It has been shown that LLM performance on tasks can vary by between 8% and 50% just by optimizing the instructions[64]. The sensitivity of LLMs to the order of presented information has been well documented on multiple choice questions [66, 43] and information retrieval[33]. The difficulty LLMs have in interpreting numbers[51] and solving simple arithmetic[17] is an active research topic[23, 52]. Even the largest models currently available, PaLM 2 and GPT4, perform poorly on instruction following tests[67]. Our analysis demonstrates how these current limitations of LLMs become harmful in medical contexts where robustness and consistency are essential. We argue that these understudied aspects of model performance should become normal parts of medical model evaluations and that all of these issues must be addressed before LLMs can be considered for clinical decision making.

## 4 Limitations

While we have been able to demonstrate the limitations of current leading large language models for clinical decision making, we consider the following limitations of our study.

First, as we are using a dataset of real-world clinical data, we rely on the accuracy of the discharge diagnosis written by the attending clinicians. Since a diagnosis using our dataset must be made with available information and without direct access to the patient, the task of coming to the exact diagnosis of the attending physician is a difficult one. We can only provide the information that was gathered during the patient’s hospital stay, so model requests for information not in the MIMIC-IV database must be denied. However, as the MIMIC-IV database contains all data gathered during a patient’s hospital stay, we can assume that all information required for a diagnosis and treatment plan is contained within our data considering the attending physicians successfully diagnosed and treated all patients. Furthermore, being flexible enough to handle acute restrictions, such as unavailable imaging modalities or laboratory tests, and still come to a correct diagnosis is a desirable ability for any real-world clinical AI application. Due to this difficulty, we were lenient in our evaluation of the diagnoses, accepting alternative names for the pathologies, as long as they were medically correct (see Supplementary Table 5).

Another issue concerns the handling of data gathered multiple times during a patient’s hospital stay, such as laboratory data. As we test LLMs in an emergency-department-centered clinical decision making scenario, we only provide the models with the first collected value for each laboratory test as they are likely most indicative for diagnostic purposes rather than treatment monitoring. Thus, we currently do not capture the changes in laboratory test values if the patient’s condition deteriorates over their hospital stay. This could be remedied by examining all time points and determining the most abnormal test result to be returned or by allowing multiple requests for laboratory tests to return successive test results. However, both of these approaches would widen the temporal gap of provided test results, possibly providing conflicting diagnostic signals. Considering LLMs also have poor temporal reasoning capabilities[62], simply including the timestamp would most likely not be an adequate solution.

Our comparison between models and clinicians included only three doctors in residency from Germany and one senior hospitalist from the United States. Increasing the diversity and number of clinicians as well as the number of patient cases evaluated would give a more nuanced view of model performance compared to practicing hospitalists. Future models could possibly soon reach or even outperform clinicians in residency and thus provide a low-cost, interactive second opinion to consult, as is already the case for AI models in other areas such as mammography screening [35].

Lastly, we only examined the initial complaint of abdominal pain and four related diagnostic endpoints in our study. While these pathologies are clinically important and well suited for our analysis, it will be important to test future models on both additional diagnostic endpoints and a broader range of initial complaints. A clinical decision making model must show strong performance across all possible pathologies of a particular initial complaint to guarantee adequate patient care without extensive preliminary diagnoses by clinicians. As each initial complaint has its own set of relevant diagnostic exams and is often best investigated with complaint-dependant exams, model performance could vary greatly. We believe the MIMIC-IV database to be a rich resource to help create such additional datasets considering its unfiltered inclusion of all patients that presented to the emergency room.

## 5 Conclusion

In conclusion, our study presents the first analysis of the capabilities of current state-of-the-art large language models on real-world data in a realistic clinical decision making scenario. Our main finding is that current models do not achieve satisfactory diagnostic accuracy, performing significantly worse than trained physicians, and do not follow treatment guidelines, thus posing a serious risk to the health of patients. This is exacerbated by the fact that they do not request the necessary exams for a safe differential diagnosis, as required by diagnostic guidelines, indicating a tendency to diagnose before fully understanding a patient’s case. Furthermore, we show LLMs are distracted by relevant diagnostic information, are sensitive to the order of diagnostic tests, and struggle to follow instructions, prohibiting their autonomous deployment in the clinic and requiring extensive clinician supervision.

By sourcing our dataset from real clinical cases and closely aligning our evaluation criteria with official diagnostic and treatment guidelines, we present the first analysis to help physicians understand how well LLMs would perform in the clinic today. While our findings cast doubt on the suitability of LLMs for clinical decision making as they currently exist, we believe there lies great potential in their use after the issues raised are resolved. By making our dataset and framework freely available we hope to guide the development of the next generation of clinical AI models and contribute towards a future where physicians can feel confident in using safe and robust models to improve patient outcomes.

## 6 Methods

### 6.1 MIMIC-CDM Dataset

We created our curated dataset of 2400 patients which we call MIMIC-Clinical Decision Making (MIMIC-CDM), using the MIMIC-IV Database [22]. The MIMIC-IV Database is a comprehensive, publicly available database of the de-identified electronic health records of almost 300,000 patients that presented to the Beth Israel Deaconess Medical Center (BIDMC) in Boston, Massachusetts from 2008 to 2019. It contains real patient cases from the hospital and includes all recorded measurements such as laboratory and microbiology test results, diagnoses, procedures, treatments and free-text clinical notes such as discharge summaries and radiologist reports.

In this work, we focus on four target pathologies for which we filter: appendicitis, cholecystitis, diverticulitis, and pancreatitis. As we are only testing for these pathologies, we must ensure that they are the primary diagnosis and reason for the patient presenting to the emergency department and not merely a secondary diagnosis during a longer and more serious hospital admission. Thus, we first filtered patients for our targets using the diagnosis table which contains all recorded diagnostic International Statistical Classification of Diseases and Related Health Problems (ICD) codes. Then, we manually checked the discharge diagnosis of each patient’s discharge summary and only included those patients whose very first primary diagnosis was one of our pathologies. If any other diagnosis was written in the discharge diagnosis before one of our targets, the patient’s case was removed from the dataset. If a patient was diagnosed with more than one of the four pathologies included in our analysis, the patient was removed from the dataset.

After filtering for the appropriate pathologies, we split the discharge summary into its individual sections, extracting the history of present illness and physical examination. First, we removed all patients where the pathology was mentioned in the history of present illness as these admissions were mostly transfers where the diagnosis had already been established and the hospital admission data was thus missing the initial emergency-department test results relevant for diagnostic purposes. Furthermore, we removed all patients where no physical examination was included as this is a crucial source of information according to the diagnostic guidelines of each pathology.

We gathered all laboratory and microbiology tests recorded during a patient’s hospital admission and those up to one day before admission. We included tests up to one day before admission as the MIMIC-IV documentation states that there are millions of laboratory tests that are not associated with any hospital admission but can be joined to patient stays using the patient’s id, recorded test time, and hospital admission time. The tests before the official start of the admission often had highly relevant values for diagnostic purposes and were thus included, though only if they were not associated with any other hospital admission. If a laboratory test was recorded multiple times, we included only the first entry in our dataset to simulate a therapy-naive diagnostic clinical-decision-making scenario. Furthermore, we saved all reference ranges of the laboratory tests provided by MIMIC and established a comprehensive dictionary mapping possible synonyms and abbreviations of tests to their original entry to be able to interpret all requests of the LLMs for test results. This dictionary of synonyms was constantly expanded during initial testing of the models until no unrecognized tests were recorded. The dictionary also contains common laboratory test panel names that map to a list of the individual tests of that panel, such as complete blood count, basic metabolic panel, liver function panel, renal function panel, and urinalysis, among others.

Similar to the laboratory data, many radiology reports were not associated with any hospital admission but their timestamp was a few hours before the recorded start of the hospital admission. These often contained diagnosis-relevant information and so we used the same 24 hour inclusion criteria as for the laboratory results and again allowed only those exams not associated with any other hospital admission. Next, we established a list of uniquely identifying keywords for each anatomical region and imaging modality. We used this list of keywords to determine the region and modality of each included report from its MIMIC-IV provided exam name. Mappings were made for special exams such as CTU to CT and MRCP to MRI, to provide them if, for example, a CT scan or MRI was requested, due to their low frequency. We also used this list when interpreting the model requests for imaging information during evaluation. We manually checked and adjusted the keywords until all reports in MIMIC-IV were correctly classified. Radiology reports were split into report sections and only the *findings* section was included. This was done as many other sections such as *conclusions* or *impressions* contained the diagnosis of the radiologist, which would have made the task trivial.

Finally, all procedures and operations performed during a patient’s hospital stay were saved to understand which patient-specific treatments were undertaken. The procedures in the MIMIC-IV procedures table saved as ICD9 and ICD10 codes were extracted and combined with the free-text procedures section from each patient’s discharge summary. The free-text extraction from the discharge summary was required as many essential procedures, including surgeries, were often not included in the procedures table.

A final round of data cleaning replaced any remaining mentions of the primary diagnosis with three underscores ‘’ which is used by MIMIC-IV to censor data such as a patient’s name or age. To increase data quality we excluded patients that had no associated laboratory tests or for which no abdominal imaging was recorded. The final dataset, MIMIC-CDM, contains 2400 unique patients presenting to the emergency department with one of the four target pathologies (957 appendicitis, 648 cholecystitis, 257 diverticulitis, 538 pancreatitis) and whose makeup is detailed in Figure 1a. The dataset contains physical examinations for all patients (2400), 138,788 laboratory results from 480 unique laboratory tests and 4403 microbiology results from 74 unique tests. Furthermore, MIMIC-CDM contains 5959 radiology reports, including 1836 abdominal CTs, 1728 chest x-rays, 1325 abdominal ultrasounds, 342 abdominal x-rays and 227 MRCPs. Finally, there were 395 unique procedures recorded over all patients, with a total of 2917 ICD procedures plus the 2400 free text procedures specified in the discharge summaries. Supplementary Table 2 shows the age, sex and race statistics of the patients in the dataset split up by pathology. As the reports provided were de-identified, the models did not have access to any of these characteristics during evaluation.

A second version of the dataset, which we call MIMIC-CDM-Full Information (MIMIC-CDM-FI), combines all the information required for diagnosing each case and presents it all at once. Here we include the history of present illness, physical examination, all abdominal imaging and all laboratory data helpful for both reaching the correct diagnosis and ruling out differential diagnoses. To determine which laboratory data to include, we used the diagnostic guidelines of each disease: appendicitis[16], cholecystitis[44], diverticulitis[25], and pancreatitis[32]. The specific tests included in each category can be found in Supplementary Table 4. The information is presented in the order: history of present illness, physical examination, laboratory results, imaging. The imaging is ordered by chart time from earliest to latest.

A subset of 80 representative patients of the MIMIC-CDM-FI dataset was randomly selected to be used for comparison with physicians. The subset is evenly split between the four target pathologies with 20 patients randomly selected from each pathology and matching the makeup of the full dataset, as shown in Supplementary Table 3. For the physicians, the data was prepared as a PDF and the information was provided exactly in the same order and quantity as for the models. Reference ranges were included when provided by MIMIC-IV. The abbreviations in the history of present illness and physical examination were replaced with unabbreviated text for the German doctors, as they were unfamiliar with US-specific abbreviations. The models performed worse with unabbreviated text (Supplementary Fig. 14). The laboratory data were provided as a table in the PDF to increase readability. To mitigate the risk of physicians recognizing the pattern of four distinct target pathologies, a further five patients each of patients presenting with gastritis, urinary tract infaction, esophageal reflux and hernia were included. Thus, the final dataset used in the reader study spanned 100 patients, 80 of which are part of MIMIC-CDM.

### 6.2 Evaluation Framework

To realistically test the capabilities of LLMs on the task of clinical decision making, we simulated a clinical environment in which a patient presents to the emergency department with acute abdominal pain and information must be iteratively gathered before a final diagnosis is made. The LLM is tasked with the Clinical Decision Making (CDM) task (Supplementary Materials B.1) which instructs it to consider a patient’s symptoms and gather information to come to a diagnosis and treatment plan while also explaining the two formats it should answer with. Both formats ask the LLM to first think (i.e. consider the evidence, which has been shown to improve the quality of reasoning and future actions[65]), and then either request more information or provide a diagnosis and treatment plan. If it chooses to request more information, it must state ‘Action:’ followed by either ‘Physical Examination’, ‘Laboratory Test’, or ‘Imaging’. Additionally, it must provide an ‘Action Input’ which specifies what information is desired from the action (i.e. ‘Complete Blood Count’ or ‘Abdominal Ultrasound’). The ‘Action Input’ field is ignored if a physical examination is requested. The second format is to be used when the model decides enough information has been gathered for a diagnosis, and asks the model to consider the evidence one last time and then provide a final diagnosis and treatment plan.

The model is initially presented with these instructions and the history of present illness of the patient and then prompted to record its ‘Thoughts:’, thus beginning the clinical decision making task. Outputs are generated until either a stop token is reached or the model generates the ‘Observation(s):’ phrase, indicating that it has reached the end of its action request and would potentially start hallucinating the result of its request. We stop model text generation here and then examine the response of the model, extracting which action was desired and what the input to that action is. If the model does not follow the instructions and, for example, writes ‘Perform a physical examination’ instead of ‘Action: Physical Examination’, we still provide the appropriate information but record every instance of it not following instructions for our evaluations (see Figure 6). We call these errors ‘Next Action Errors’.

If the requested information is available for that patient case, we return it and prompt the model again to consider the evidence. If the information is not available, we inform the model and ask for an alternative action. We return only the laboratory tests and radiologist reports that were specifically requested. Laboratory tests are compared to the previously mentioned dictionary of available tests to return the best match. If no match is found, ‘N/A’ is returned. Requests for imaging have the exam modality and anatomical region extracted using the aforementioned keyword lists (see Materials Section 6.1) and used to match against those saved for that patient in MIMIC-CDM. If multiple reports exist for a modality and region combination, we return the first report chronologically. The next request for an imaging examination of that modality and region will return the next report chronologically. Once there are no reports left to return, we inform the model that we can no longer provide reports of that modality and region combination. If a physical examination was requested, we return the entire physical examination regardless of any specifications made in the ‘action input’ field. We do this because we consider it best practice to perform a complete physical examination of a patient rather than only partially and reliably separating a physical examination report into its parts is difficult due to their free-form and heavily abbreviated style. If an invalid tool is requested (‘hallucinated’), we state that the tool does not exist and remind it which tools are available, or that it should make a diagnosis and provide a treatment plan. These occasions are also recorded as tool hallucinations for our evaluations (see Figure 6). An example exchange between an LLM and our framework can be seen in Figure 1b and Supplementary Section C.

We repeat this process, prompting the LLM to think and in turn receiving requests for information. Once the model decides that it has gathered sufficient information, it outputs its final diagnosis and treatment plan, ending the clinical decision making task for that patient. The final diagnosis is then evaluated to see if it contains the recorded pathology of the patient. In addition to a direct match of the pathology name (i.e. appendicitis, cholecystitis, diverticulitis, pancreatitis), we allow for a range of alternative phrasings as long as they are medically correct (see Supplementary Table 5). If multiple diagnoses are given, we only examine the first diagnosis mentioned. This is how we calculate the diagnostic accuracy for all analyses. A new instance of the task is then started for the next patient.

As LLMs can only take a limited amount of words as input, we monitor the length of the conversation and if we approach the input limit of the model, we ask it to summarize the information it has received so far to reduce the length of the conversation (Supplementary Materials B.2). We first summarize each gathered piece of information individually, leaving the initial history of present illness and instructions untouched. As LLMs have no memory and interpret each request independently, we replace the original pieces of information with the summaries. If we have summarized all steps of the interaction and still approach the limit of the model, we force a generation of diagnosis and treatment plan.

For the MIMIC-CDM-FI dataset we instruct the model to consider the facts of the case and then provide a diagnosis and only a diagnosis (Supplementary Materials B.3). As explained in Materials Section 6.1, the MIMIC-CDM-FI dataset includes the history of present illness, physical examination, all relevant laboratory results and every radiologist report where the abdominal region was inspected. Radiologist reports of other regions were not included due to the input length limits of the models. If including all of this information exceeds the input length of the model, we ask the LLM to summarize each radiologist report individually. If the input length is still exceeded, we ask the LLM to summarize all imaging information at once. In the rare cases where the input length continues to be exceeded, we remove words from the final imaging summary until there is enough space for a diagnosis (i.e. 25 tokens or 20 words).

### 6.3 Models

An overview of the models included and considered is given in Table 1.

**Table 1:** An overview of the considered LLMs and their properties. Due to the data usage agreement of MIMIC-IV, only open access models that can be downloaded can be used with the data. Thus, only LLMs based on Llama 2 were used in this study. * Meta defines ‘public data’ as a ‘mix of data from publicly available sources’. *†* No further information provided. indicates no information has been made public.

| **Model** | **Base** | **Parameters** | **Training Dataset** | **Downloadable** |
|---|---|---|---|---|
| Llama 2 Chat[58] | Llama 2[58] | 70B | Public data* | ✓ |
| OASST[30] | Llama 2[58] | 70B | Public data*<br>Open Source Data | ✓ |
| WizardLM[63] | Llama 2[58] | 70B | Public data*<br>Evol-Instruct Generated[63] | ✓ |
| Clinical Camel[56] | Llama 2[58] | 70B | Public data*<br>ShareGPT<br>PubMed Articles (pre 2021)[56]<br>MedQA[27] | ✓ |
| Meditron[12] | Llama 2[58] | 70B | Public data*<br>Clinical Guidelines<br>Public PubMed Abstracts[12]<br>Public PubMed Papers[12]<br>RedPajama[14] | ✓ |
| Chat-GPT[41] | GPT3.5[9] | ??? | User Conversations†<br>Common Crawl[47]<br>WebText2[45]<br>Books1[29]<br>Books2[29]<br>Wikipedia | ✗ |
| GPT4[40] | ??? | ??? | ??? | ✗ |
| Med-PaLM[49] | Flan-PaLM[13] | 540B | Webpages†<br>Wikipedia†<br>Social Media†<br>Github†<br>News Articles†<br>Books†<br>473 Instruction fine-tuning Datasets[13]<br>HealthSearchQA[49]<br>MedicationQA[2]<br>LiveQA[1] | ✗ |
| Med-PaLM 2[50] | PaLM 2[4] | 340B | Web Documents†<br>Books†<br>Code†<br>Mathematics†<br>Conversational Data†<br>MedQA[27]<br>HealthSearchQA[49]<br>MedicationQA[2]<br>LiveQA[1] | ✗ |

When deciding which models to test, we started by only considering models with a context length of 4096 tokens due to the large amounts of text contained within the MIMIC-CDM clinical cases. The context length defines how many combined tokens a large language model can read and write. For example, if a model has a context length of 2048 and receives an input of 2000 tokens, it can only generate 48 new tokens. A minimum length of 4096 is required, as the average number of tokens of relevant information per patient case in MIMIC-CDM-FI is 2080 tokens with a maximum count of 15,023 tokens. If one considers the extra tokens that are required for the information gathering back-and-forth using MIMIC-CDM data, this quickly exceeds the limit of 2048 tokens of smaller models (context length windows almost always differ in powers of 2).

Next, we considered which open access models performed best on medical reasoning tasks. To gauge model strength, we used the MedQA[27] dataset as it is comprised of 12,723 questions from the USMLE and is thus a good gauge of general medical knowledge contained within the model. As of the time of writing, Llama 2 is the leading open-access model on the MedQA (USMLE) dataset, with the 70B model achieving a score of 58.4[12] exceeding that of GPT3.5 which scored only 53.6[38].

To effectively complete the clinical decision making task without specific fine-tuning to the task and format, the model must be instruction fine-tuned. Instruction fine-tuning involves training a model to adapt to a wide range of new tasks so that it can, with minimal instruction or example, complete an unseen task, like our clinical decision making objective. The most popular and performant instruction fine-tuned versions of Llama 2 are Llama 2 Chat, fine-tuned by Meta themselves; WizardLM, fine-tuned by Microsoft using evolutionary algorithm (Evo-Instruct) generated training data; and OASST, fine-tuned using a crowd-sourced collection of 161,443 messages. Currently the only two existing medically fine-tuned versions of Llama 2 with a context length of 4096 and 70B parameters are Clinical Camel and Meditron. Neither has been extensively instruction fine-tuned and thus they both generated nonsensical and repetitive responses to the clinical decision making objective using MIMIC-CDM data.

Currently, the most popular and leading closed-source LLMs for medical question answering are Chat-GPT (MedQA: *∼*53.6[38]), GPT-4 (MedQA: 90.2[39]), Med-PaLM (MedQA: 67.2[50]), and Med-PaLM 2 (MedQA: 86.5[50]). As previously stated, due to the signed data usage agreements of the MIMIC-IV database, the data cannot be sent to external servers[19], precluding its use with closed-source models that are only accessible through an API and whose models cannot be downloaded.

Furthermore, Chat-GPT is fine-tuned primarily through user conversations with the model, and since it is impossible to know if portions of the MIMIC-IV database have already been used for queries by users less aware of the data usage agreement[19], the data could already have been seen by the model during training, invalidating any results it produces. Little to no information is known about the training data of GPT4, giving rise to analogous concerns about its performance. While the exact pre-training data of Llama 2 is also not known, Meta has stated that it only used ‘public available online data’, which strongly mitigates the risk of MIMIC-IV data having being used. Med-PaLM and Med-PaLM 2 achieve strong scores on MedQA but the exact data used for training are unknown, the models are only accessible through an API, and access to the models is currently unavailable for all researchers. Repeated requests for access were denied.

We strongly agree with current sentiment that open source models must drive progress in the field of medical AI due to patient privacy and safety concerns, corporate lack of transparency, and the danger of unreliable external providers[57]. It is a serious risk to patient safety if key medical infrastructure is based on external company APIs and models whose performance could change erratically with updates and which could generally be deactivated for any reason.

For each model tested, we downloaded and used the GPTQ quantized version from huggingface, the central repository for all LLM models. GPTQ quantization reduces the numerical precision of the weights while monitoring the generated output to reduce the GPU memory requirements of a model while not significantly degrading performance[20]. The GPTQ parameters of the downloaded models were 4 bits, 32 group size, act order true, 0.1 damp% and 4096 sequence length. This gives the highest possible inference quality while reducing model size to around 40 GB which can fit onto a single A40 GPU. This reflects an economically realistic scenario of a single high-end GPU being used to host the model to run the clinical decision making task. A fixed seed of 2023 and greedy decoding were used for all experiments making all results deterministic and reproducible, except for the evaluation on the 80 patient subset for comparison with clinicians where 10 different seeds were used for increased statistical power.

### 6.4 Data and Code Availability

The dataset is available to all researchers who create an account on https://physionet.org/ and follow the steps to gain access to the MIMIC-IV database (https://physionet.org/content/mimiciv/2.2/). Access is given after completing the “CITI Data or Specimens Only Research” training course. The data use agreement of physionet for “credentialed health data” must also be signed. The dataset can then be recreated using the code found at: https://github.com/paulhager/MIMIC-Clinical-Decision-Making-Dataset. The code to create the dataset uses python v3.10 and pandas v2.1.3.

The publication of the dataset on the Physionet website for those with access to MIMIC is currently under review. Once it has been accepted, anyone credentialed to access MIMIC will be able to download the data directly.

The evaluation framework used for this study can be found at: https://github.com/paulhager/MIMIC-Clinical-Decision-Making-Framework. The analysis framework to evaluate all results, generate all plots and do all statistical analysis can be found at: https://github.com/paulhager/MIMIC-Clinical-Decision-Making-Analysis. All code uses python v3.10, pytorch v2.1.1, transformers v4.35.2, spacy v3.4.4, langchain v0.0.339, optimum v1.14, thefuzz v0.20, exllamav2 v0.0.8, nltk v3.8.1, negspacy v1.0.4, scispacy v0.5.2

### 6.5 Statistics

All statistical tests were conducted using the Python programming language, version 3.10 and using the SciPy library. Comparisons of means were tested for statistical significance using two-sided Student’s *t*-tests with unequal variances (tested through Bartlett’s tests). To account for multiple comparisons, *p*-values were Bonferroni corrected with a multiplier of 5 for the comparison of the doctors against the models and 3 for the comparison of the specialist and generalist models.

## Data Availability

The dataset is available to all researchers who create an account on https://physionet.org/ and follow the steps to gain access to the MIMIC-IV database (https://physionet.org/content/mimiciv/2.2/). Access is given after completing the "CITI Data or Specimens Only Research" training course. The data use agreement of physionet for "credentialed health data" must also be signed. The dataset can then be recreated using the code found at: https://github.com/paulhager/MIMIC-Clinical-Decision-Making-Dataset. The code to create the dataset uses python v3.10 and pandas v2.1.3.

## A Dataset Statistics

**Table 2:**
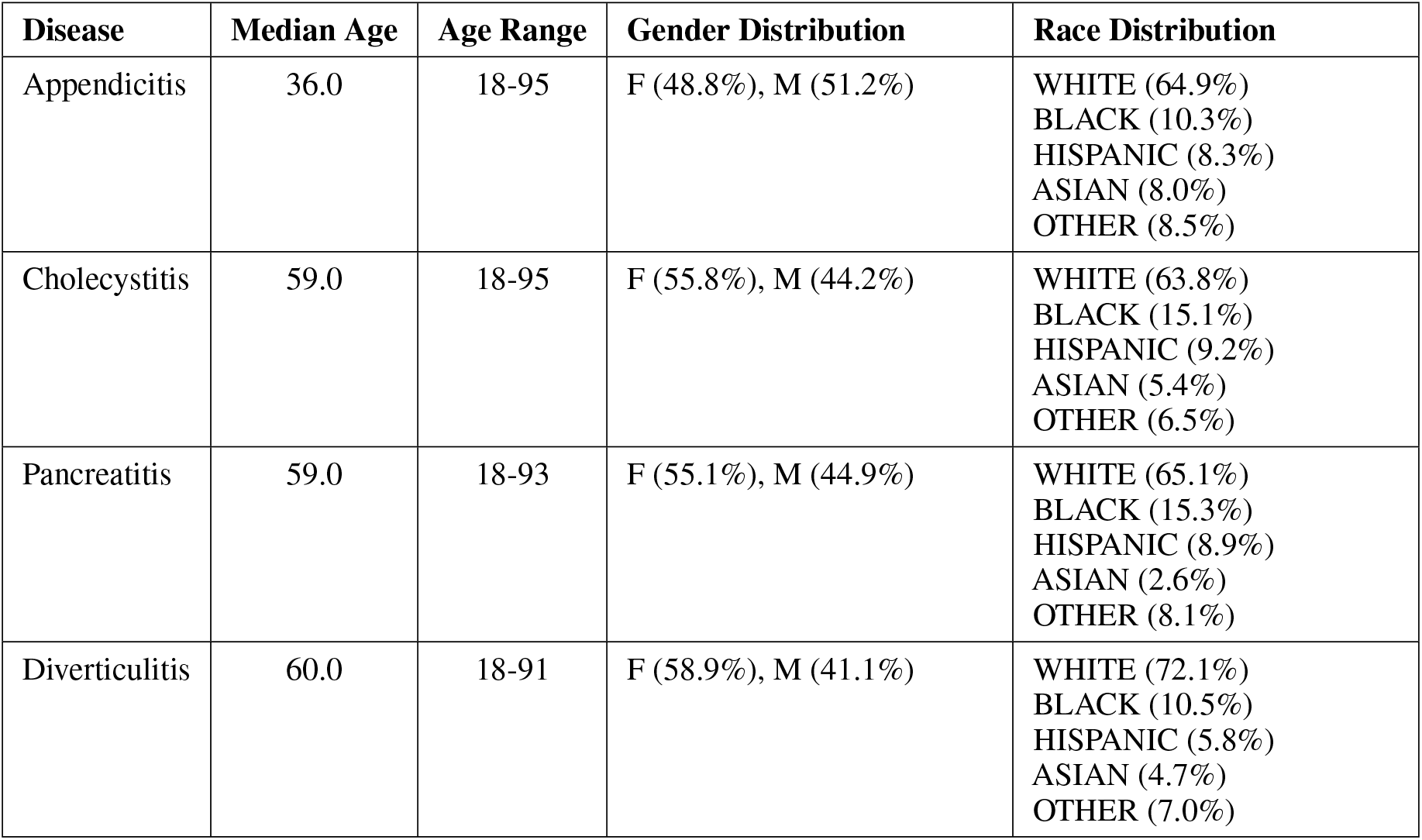
Demographic Statistics of Patients with Different Diseases.

**Table 3:** Demographic Statistics of Physician Comparison Subset of 80 patients.

| **Disease** | **Median Age** | **Age Range** | **Gender Distribution** | **Race Distribution** |
|---|---|---|---|---|
| Appendicitis | 38.5 | 20-75 | F (45.0%), M (55.0%) | WHITE (65.0%)<br>BLACK (15.0%)<br>HISPANIC (5.0%)<br>ASIAN (5.0%)<br>OTHER (10.0%) |
| Cholecystitis | 66.5 | 31-85 | F (45.0%), M (55.0%) | WHITE (65.0%)<br>BLACK (10.0%)<br>HISPANIC (5.0%)<br>ASIAN (5.0%)<br>OTHER (15.0%) |
| Pancreatitis | 52.5 | 20-82 | F (60.0%), M (40.0%) | WHITE (50.0%)<br>BLACK (10.0%)<br>HISPANIC (15.0%)<br>ASIAN (0.0%)<br>OTHER (25.0%) |
| Diverticulitis | 56.0 | 31-85 | F (45.0%), M (55.0%) | WHITE (75.0%)<br>BLACK (15.0%)<br>HISPANIC (0.0%)<br>ASIAN (5.0%)<br>OTHER (5.0%) |

## B Prompts

### B.1 CDM Template

~~~
{system_tag_start}You are a medical artificial intelligence assistant. You give helpful, detailed and factually correct answers to the doctors questions to help him in his clinical duties. Your goal is to correctly diagnose the patient and provide treatment advice. You will consider information about a patient and provide a final diagnosis.
~~~

~~~
You can only respond with a single complete Thought:
~~~

~~~
Action:
~~~

~~~
Action Input:
~~~

~~~
format OR a single
~~~

~~~
Thought:
~~~

~~~
Final Diagnosis:
~~~

~~~
Treatment:
~~~

~~~
format. Keep all reasoning in the Thought section. The Action, Action Input, Final Diagnosis, and Treatment sections should be direct and to the point. The results of
~~~

~~~
the action will be returned directly after the Action Input field in the “Observation:” field.
~~~

~~~
Format 1:
~~~

~~~
Thought: (reflect on your progress and decide what to do next) Action: (the action name, should be one of [{tool_names}]) Action Input: (the input string to the action)
~~~

~~~
Observation: (the observation from the action will be returned here)
~~~

~~~
OR
~~~

~~~
Format 2:
~~~

~~~
Thought: (reflect on the gathered information and explain the reasoning for the final diagnosis)
~~~

~~~
Final Diagnosis: (the final diagnosis to the original case) Treatment: (the treatment for the given diagnosis)
~~~

~~~
The tools you can use are:
~~~

~~~
Physical Examination: Perform physical examination of patient and receive the observations.
~~~

~~~
Laboratory Tests: Run specific laboratory tests and receive their values. The specific tests must be specified in the ’Action Input’ field.
~~~

~~~
Imaging: Do specific imaging scans and receive the radiologist report.
~~~

~~~
Scan region AND modality must be specified in the ’Action Input’ field.{add_tool_descr}{system_tag_end}{user_tag_start}{examples}Consider the following case and come to a final diagnosis and treatment by thinking, planning, and using the aforementioned tools and format.
~~~

~~~
Patient History:
~~~

~~~
{input}{user_tag_end}{ai_tag_start}Thought:{agent_scratchpad}
~~~

### B.2 CDM Observation Summarize Template

~~~
{system_tag_start}You are a medical artificial intelligence assistant. Your goal is to effectively, efficiently and accurately reduce text without inventing information. You want to return verbatim observations that are abnormal and of interest to a possible diagnosis of the patient. Normal observations can be combined. Do not invent information. Use medical abbreviations when possible to save characters. Put the most important information first.{system_tag_end}{user_tag_start}Please summarize the following result:
~~~

~~~
{observation}{user_tag_end}{ai_tag_start} Summary:
~~~

### B.3 CDM-FI Template

~~~
{system_tag_start}You are a medical artificial intelligence assistant. You directly diagnose patients based on the provided information to assist a doctor in his clinical duties. Your goal is to correctly diagnose the patient. Based on the provided information you will provide a final diagnosis of the most severe pathology. Don’t write any further information. Give only a single diagnosis.{system_tag_end}{fewshot_examples}{user_tag_start}Provide the most likely final diagnosis of the following patient.
~~~

~~~
{input}{diagnostic_criteria}{user_tag_end}{ai_tag_start}Final Diagnosis:
~~~

### B.4 Reference Range Test Zeroshot Template

~~~
{system_tag_start}You are a technical AI assistant working in a laboratory that handles tests for a hospital. You are good at interpreting numbers. You are responsible for reviewing the results of lab tests and determining whether they are Low, Normal, or High. You will be given the test, its value and then the reference range for that test, which will be written as “Reference Range [Lower Reference Range – Upper Reference Range]”. You will write just one word, indicating if the test results are Low, Normal, or High. Do not write anything other than your one word answer.{system_tag_end}{user_tag_start}{lab_test_string_rr}{user_tag_end}{ai_tag_start}Test Result:
~~~

### B.5 CDM-FI No Final Template

~~~
{system_tag_start}You are a medical artificial intelligence assistant. You directly diagnose patients based on the provided information to assist a doctor in his clinical duties. Your goal is to correctly diagnose the patient. Based on the provided information you will provide the diagnosis. Don’t write any further information. Give only a single diagnosis.{system_tag_end}{fewshot_examples}{user_tag_start}Provide the diagnosis of the following patient.
~~~

~~~
{input}{diagnostic_criteria}{user_tag_end}{ai_tag_start}Diagnosis:““”
~~~

### B.6 CDM-FI Main Diagnosis Template

~~~
{system_tag_start}You are a medical artificial intelligence assistant. You directly diagnose patients based on the provided information to assist a doctor in his clinical duties. Your goal is to correctly diagnose the patient. Based on the provided information you will provide the main diagnosis. Don’t write any further information. Give only a single diagnosis.{system_tag_end}{fewshot_examples}{user_tag_start}Provide the main diagnosis of the following patient.
~~~

~~~
{input}{diagnostic_criteria}{user_tag_end}{ai_tag_start}Main Diagnosis:”“”
~~~

### B.7 CDM-FI Primary Diagnosis Template

~~~
{system_tag_start}You are a medical artificial intelligence assistant. You directly diagnose patients based on the provided information to assist a doctor in his clinical duties. Your goal is to correctly diagnose the patient. Based on the provided information you will provide the primary diagnosis. Don’t write any further information. Give only a single diagnosis.{system_tag_end}{fewshot_examples}{user_tag_start}Provide the primary diagnosis of the following patient.
~~~

~~~
{input}{diagnostic_criteria}{user_tag_end}{ai_tag_start}Primary Diagnosis:““”
~~~

### B.8 CDM-FI No System Template

~~~
{system_tag_start}{system_tag_end}{fewshot_examples}{user_tag_start}Provide the most likely final diagnosis of the following patient.
~~~

~~~
{input}{diagnostic_criteria}{user_tag_end}{ai_tag_start}Final Diagnosis:““”
~~~

### B.9 CDM-FI No User Template

~~~
{system_tag_start}You are a medical artificial intelligence assistant. You directly diagnose patients based on the provided information to assist a doctor in his clinical duties. Your goal is to correctly diagnose the patient. Based on the provided information you will provide a final diagnosis of the most severe pathology. Don’t write any further information. Give only a single diagnosis. {system_tag_end}{fewshot_examples}{user_tag_start}{input}{diagnostic_criteria} {user_tag_end}{ai_tag_start}Final Diagnosis:““”
~~~

### B.10 CDM-FI No Medical Template

~~~
{system_tag_start}You are an artificial intelligence assistant. You answer questions to the best of your abilities. Think hard about the following problem and then provide an answer.{system_tag_end}{fewshot_examples}{user_tag_start}Provide the most likely final diagnosis of the following patient.
~~~

~~~
{input}{diagnostic_criteria}{user_tag_end}{ai_tag_start}Final Diagnosis:““”
~~~

### B.11 CDM-FI Serious Final Template

~~~
{system_tag_start}You are a medical artificial intelligence assistant. You directly diagnose patients based on the provided information to assist a doctor in his clinical duties. Your goal is to correctly diagnose the patient. Based on the provided information you will provide a final diagnosis of the most severe pathology. Don’t write any further information. Give only a single diagnosis.{system_tag_end}{fewshot_examples}{user_tag_start}Provide the most serious final diagnosis of the following patient.
~~~

~~~
{input}{diagnostic_criteria}{user_tag_end}{ai_tag_start}Final Diagnosis:““”
~~~

### B.12 CDM-FI Minimal System Template

~~~
{system_tag_start}You are a medical artificial intelligence assistant. You diagnose patients based on the provided information to assist a doctor in his clinical duties.{system_tag_end}{fewshot_examples}{user_tag_start}Provide the most likely final diagnosis of the following patient.
~~~

~~~
{input}{diagnostic_criteria}{user_tag_end}{ai_tag_start}Final Diagnosis:““”
~~~

### B.13 CDM-FI No System No User Template

~~~
{system_tag_start}{system_tag_end}{fewshot_examples}{user_tag_start}{input} {diagnostic_criteria}{user_tag_end}{ai_tag_start}Final Diagnosis:““”
~~~

### B.14 CDM-FI No Diagnosis Prompt Template

~~~
{system_tag_start}You are a medical artificial intelligence assistant. You directly diagnose patients based on the provided information to assist a doctor in his clinical duties. Your goal is to correctly diagnose the patient. Based on the provided information you will provide a final diagnosis of the most severe pathology. Don’t write any further information. Give only a single diagnosis.{system_tag_end}{fewshot_examples}{user_tag_start}Provide the most likely final diagnosis of the following patient.
~~~

~~~
{input}{diagnostic_criteria}{user_tag_end}{ai_tag_start}““”
~~~

## C Example Exchange using Synthetic Data

In this section we show an example exchange between our simulated clinical environment provided by our evaluation framework using the MIMIC-CDM dataset and the WizardLM model. The first prompt is the system prompt given by the framework to the model explaining the task and its options. The rest of the exchange pulls (synthetic) data from MIMIC-CDM and records the LLM’s response. Seen in this exchange is an example of the model requesting an imaging modality that is not available in the dataset and also hallucinating a surgery tool that does not exist. Upon reminding it of the available tools it correctly diagnoses the patient. This example exchange uses synthetic data to further protect patient privacy.

### Evaluation Framework using MIMIC-CDM

You are a medical artificial intelligence assistant. You give helpful, detailed and factually correct answers to the doctors questions to help him in his clinical duties. Your goal is to correctly diagnose the patient and provide treatment advice. You will consider information about a patient and provide a final diagnosis.

You can only respond with a single complete Thought:

Action:

Action Input:

format OR a single Thought:

Final Diagnosis:

Treatment:

format. Keep all reasoning in the Thought section. The Action, Action Input, Final Diagnosis, and Treatment sections should be direct and to the point. The results of the action will be returned directly after the Action Input field in the “Observation:” field.

Format 1:

Thought: (reflect on your progress and decide what to do next)

Action: (the action name, should be one of [Physical Examination, Laboratory Tests, Imaging]) Action Input: (the input string to the action)

Observation: (the observation from the action will be returned here)

OR

Format 2:

Thought: (reflect on the gathered information and explain the reasoning for the final diagnosis) Final Diagnosis: (the final diagnosis to the original case)

Treatment: (the treatment for the given diagnosis) The tools you can use are:

Physical Examination: Perform physical examination of patient and receive the observations.

Laboratory Tests: Run specific laboratory tests and receive their values. The specific tests must be specified in the ‘Action Input’ field.

Imaging: Do specific imaging scans and receive the radiologist report. Scan region AND modality must be specified in the ‘Action Input’ field. Consider the following case and come to a final diagnosis and treatment by thinking, planning, and using the aforementioned tools and format.

### Evaluation Framework using MIMIC-CDM

Patient History:

___ presenting with 3 days since acute onset of pain in RLQ. Pain started ___ afternoon but worsened significantly overnight. Intense episodes woke her from sleep. Patient proceeds to experience nausea and chills. No vomiting. Patient endorses anorexia. Past Medical History: Anorexia, Anxiety. Family History:

Non-contributory.

### LLM

Thought: Given the acute onset of right lower quadrant pain, along with anorexia and right-sided flank pain, there could be several possibilities such as appendicitis, kidney stones, or inflammatory bowel disease like Crohn’s disease. However, since the patient does not have any past medical history or family history of these conditions, it might be less likely. Therefore, we need to perform further investigations to determine the exact cause of the pain.

Action: Physical Examination

Action Input: N/A

### Evaluation Framework using MIMIC-CDM

Observation: Physical Examination:

Temp: 97.8 HR: 44 BP: 104/69 RR: 17 100% Ra Gen: NAD HEENT: No scleral icterus, mucus membranes moist CV: RRR no m,r,g RESP: CTABL Abd: soft, tender to palpation inRLQ, non distended, no rebound or guarding Ext: wwpx4, palpable distal pulses

### LLM

Thought: After performing the physical examination, the vital signs appear normal except for a slightly elevated blood pressure. Findings of tenderness to palpation in the right lower quadrant of the abdomen supports the forementioned possibilities of the patient’s complaints, such as appendicitis, kidney stones, urinary tract infection, ovarial cysts or inflammatory bowel disease. We need to investigate further to rule out potential causes.

Action: Laboratory Tests

Action Input: Complete Blood Count (CBC), Liver Function Tests (LFTs), Renal panel, Urinalysis

### Evaluation Framework using MIMIC-CDM

Observation: Laboratory Tests:

(Blood) Red Blood Cells: 4.39 m/uL

(Blood) White Blood Cells: 16.8 K/uL

(Blood) Hemoglobin: 12.4 g/dL

(Blood) Hematocrit: 38.8 %

(Blood) MCV: 85.4 fL

(Blood) MCH: 27.3 pg

(Blood) MCHC: 33.4 %

(Blood) Platelet Count: 226.8 K/uL

(Blood) Lymphocytes: 22.0 %

(Blood) Basophils: 0.5 %

(Blood) Eosinophils: 1.9 %

(Blood) Monocytes: 5.5 %

(Blood) Neutrophils: 69.8 %

(Blood) RDW: 13.0 %

(Blood) Alanine Aminotransferase (ALT): 18.1 IU/L

(Blood) Asparate Aminotransferase (AST): 29.1 IU/L

(Blood) Alkaline Phosphatase: 68.8 IU/L

(Blood) Bilirubin, Total: 0.3 mg/dL (Blood) PT: 10.5 sec

(Blood) INR(PT): 0.9

(Blood) Albumin: 5.2 g/dL

(Blood) Creatinine: 0.6 mg/dl

(Urine) Ketones: NEG

(Urine) Urine Color: Yellow.

(Urine) Urine Appearance: Clear.

(Urine) Specific Gravity: 1.0

(Urine) Protein: NEG.

(Urine) pH: 5.8 units

(Urine) Bilirubin: NEG.

(Urine) Glucose: NEG.

(Urine) Urobilinogen: NEG.

(Urine) Ketone: TR.

(Urine) Nitrite: NEG.

(Urine) Leukocytes: NEG.

### LLM

Thought: The laboratory test results show normal values for most parameters analyzed. However, the patients has elevated white blood cells as a marker for inflammation. Based on the diagnostic test results so far, the patient is suspected to have an inflammation in the abdomen, such as appendicitis or inflammatory bowel disease.

Action: Imaging

Action Input: Ultrasound of the Right Lower Quadrant

### Evaluation Framework using MIMIC-CDM

Observation: Imaging:

Abdomen Ultrasound: Not available. Try a different imaging modality

### LLM

Thought: Since ultrasound is not suitable for this case, we should proceed with another imaging modality. Given the possibility of hepatobiliary involvement, a computed tomography (CT) scan would be more appropriate to visualize the internal organs better.

Action: Imaging

Action Input: Computed Tomography (CT) scan of the Abdom

### Evaluation Framework using MIMIC-CDM

Observation: Imaging:

Abdomen CT: EXAMINATION:

CT ABD AND PELVIS WITH CONTRAST:

TECHNIQUE:

MDCT imaging, extending from the lung bases to the pubic symphysis, was performed post-administration of 130cc intravenous contrast. Axial scans were analyzed alongside coronal and sagittal reformatted images.

FINDINGS:

CHEST:

Lung bases are clear without pleural or pericardial effusions.

ABDOMEN:

Liver and gallbladder present normally; portal and hepatic veins are patent. Pancreas and spleen are normal; bilateral adrenal glands are unremarkable. Kidneys show symmetrical enhancement without hydronephrosis.

Stomach and small bowel maintain normal caliber. A hyperdense, dilated structure, likely the appendix, arises from the cecal base, measuring up to 10 mm and exhibiting mucosal hyperenhancement compared to adjacent bowel. Predominant features include hyperenhancement and wall thickening, with less emphasis on fluid content. Inflammatory changes are evident at the cecal base, around the appendix, especially at its indistinct tip.

Adjacent to the ascending colon’s mesentery, two enlarged lymph nodes, 1.5 cm and 1.9 cm in size, show brisk enhancement. The smaller node contains central low-density material. Both nodes are surrounded by marked fat stranding and suspected necrotic or suppurative changes.

The ascending, transverse, and descending colon appear normal. No mesenteric free air is observed.

PELVIS:

Urinary bladder and adnexa are unremarkable. Uterus contains an intrauterine device in standard position. A small volume of simple free fluid is present in the pelvis, likely physiologic.

VESSELS:

Aorta and its major branches are normal in caliber and patent.

OSSEOUS STRUCTURES:

No focal lytic or sclerotic lesions suggestive of malignancy are identified.

### LLM

Thought: The CT scan supports the diagnosis of acute appendicitis due to the enlarged and inflamed appendix

observed. I will recommend surgical intervention for removal of the affected organ.

Action: Surgery

Action Input: Appendectomy

### Evaluation Framework using MIMIC-CDM

Observation: Provide a diagnosis and treatment OR a valid tool. That is not a valid tool, try one of [Physical Examination, Laboratory Tests, Imaging].

### LLM

Thought:

The CT scan revealed an inflamed and enlarged appendix, indicating acute appendicitis as the most likely cause of the patient’s pain. Additionally, there are two enlarged and briskly enhancing lymph nodes in the mesentery, which could suggest a secondary infectious process or inflammatory condition. The presence of a small amount of simple free fluid in the pelvis might be due to the inflammatory response.

Final Diagnosis: Acute Appendicitis

Treatment: Surgical intervention – Appendectomy

## D Laboratory Test Categories

**Table 4:** The individual laboratory test category items defined using the guidelines for appendicitis[16], cholecystitis[44], diverticulitis [25], and pancreatitis[32]. At least one of these tests must have been ordered either individually or as part of a panel for the category to be counted.

| **Pathologies** | **Category** | **Items** |
|---|---|---|
| Appendicitis<br>Cholecystitis<br>Diverticulitis<br>Pancreatitis | Inflammation | White Blood Cells<br>WBC Count<br>C-Reactive Protein<br>High-Sensitivity CRP |
| Cholecystitis | Gallbladder | Bilirubin<br>Gamma Glutamyltransferase |
| Cholecystitis | Liver | Alanine Aminotransferase (ALT)<br>Asparate Aminotransferase (AST) |
| Pancreatitis | Pancreas | Amylase<br>Lipase |
| Pancreatitis | Pancreatitis Severity | Hematocrit<br>Urea Nitrogen<br>Triglycerides<br>Calcium<br>Potassium |

## E Diagnosis Definitions

**Table 5:**
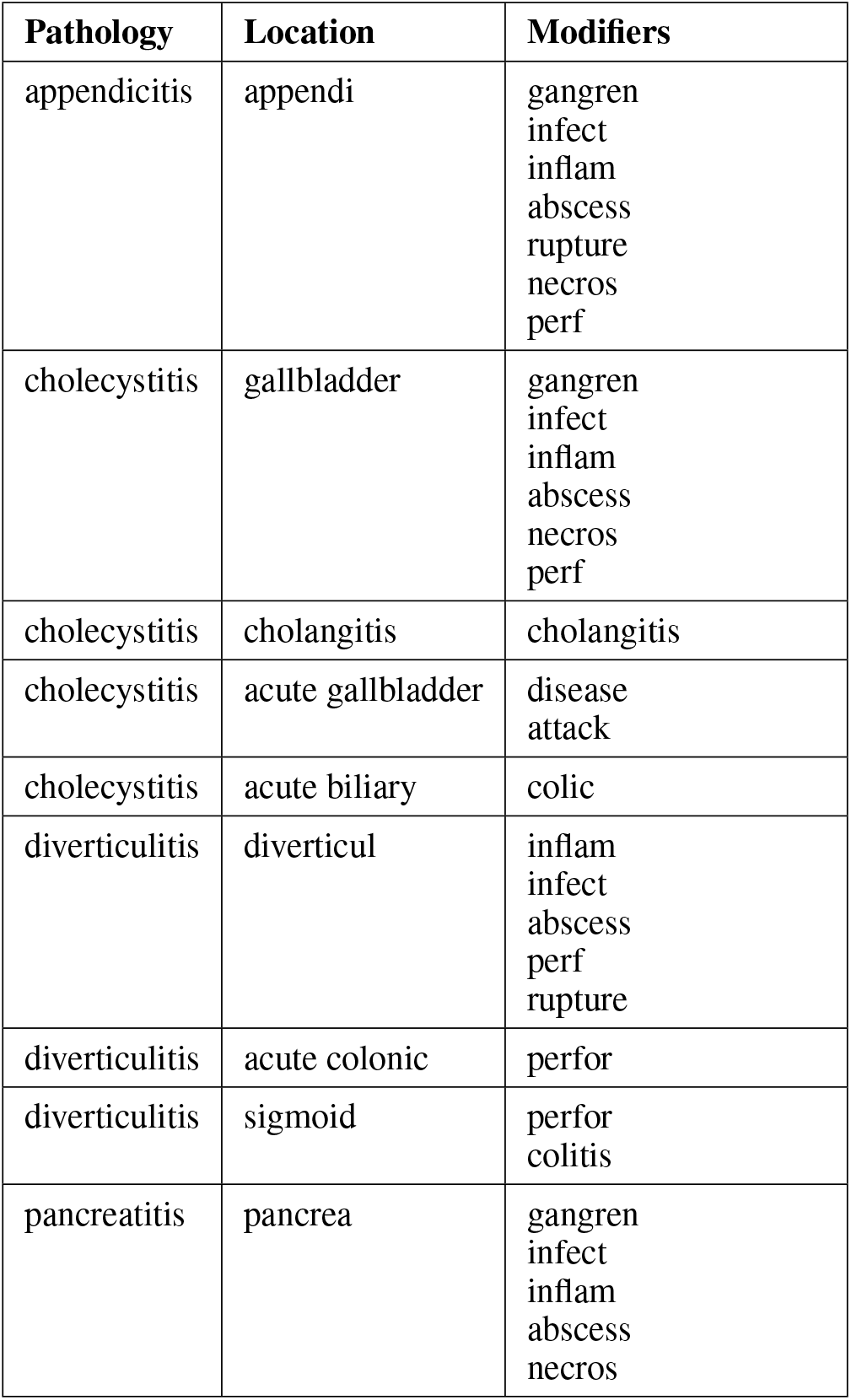
For each pathology, alternative diagnostic phrasings were accepted. If within one diagnostic phrase (delineated through e.g. periods, commas, ‘vs’, or other separators) the location and modifier occurred without negation, the diagnosis was marked as correct. For example, for cholecystitis a diagnosis of ‘Perforated gallbladder’ or ‘Gallbladder infection’ were both accepted. A diagnosis of ‘Gallbladder disease vs perforated appendix’ would not be accepted. Shortened substrings of the locations and modifiers were used to allow for alternative endings to match such as both ‘An *infect* of the gallbladder’ and ‘*Infect*ed gallbladder’, or ‘Peri*appendi*cular abscess’ and ‘Perforated *appendi*x’. Manual controls were done to verify the specificity and sensitivity of the definitions.

## F Adherence to Diagnostic Guidelines

**Figure 11:**
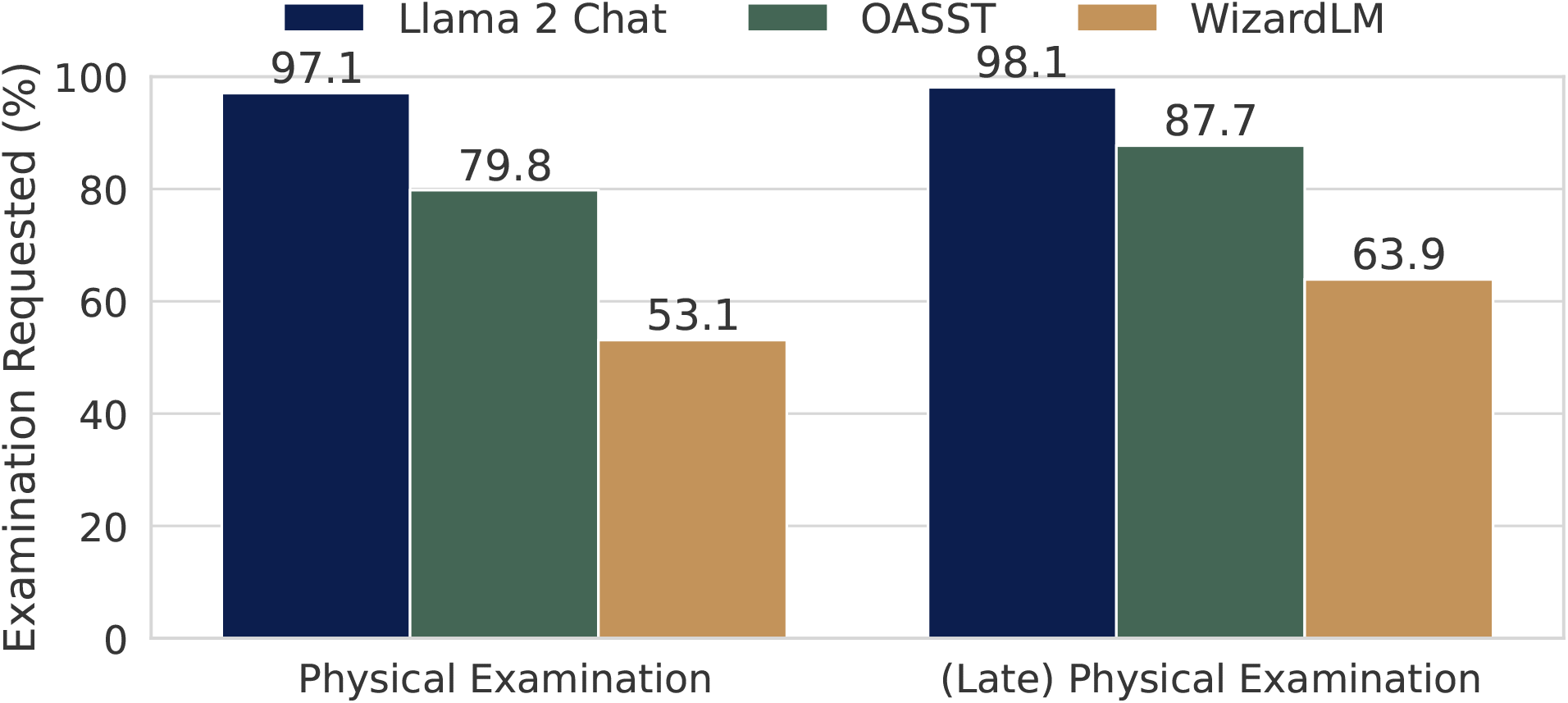
Llama 2 Chat was the only LLM that consistently requested physical examinations. (Late) Physical Examinations counted the physical examination if it wasn’t the first information requested.

**Figure 12:**
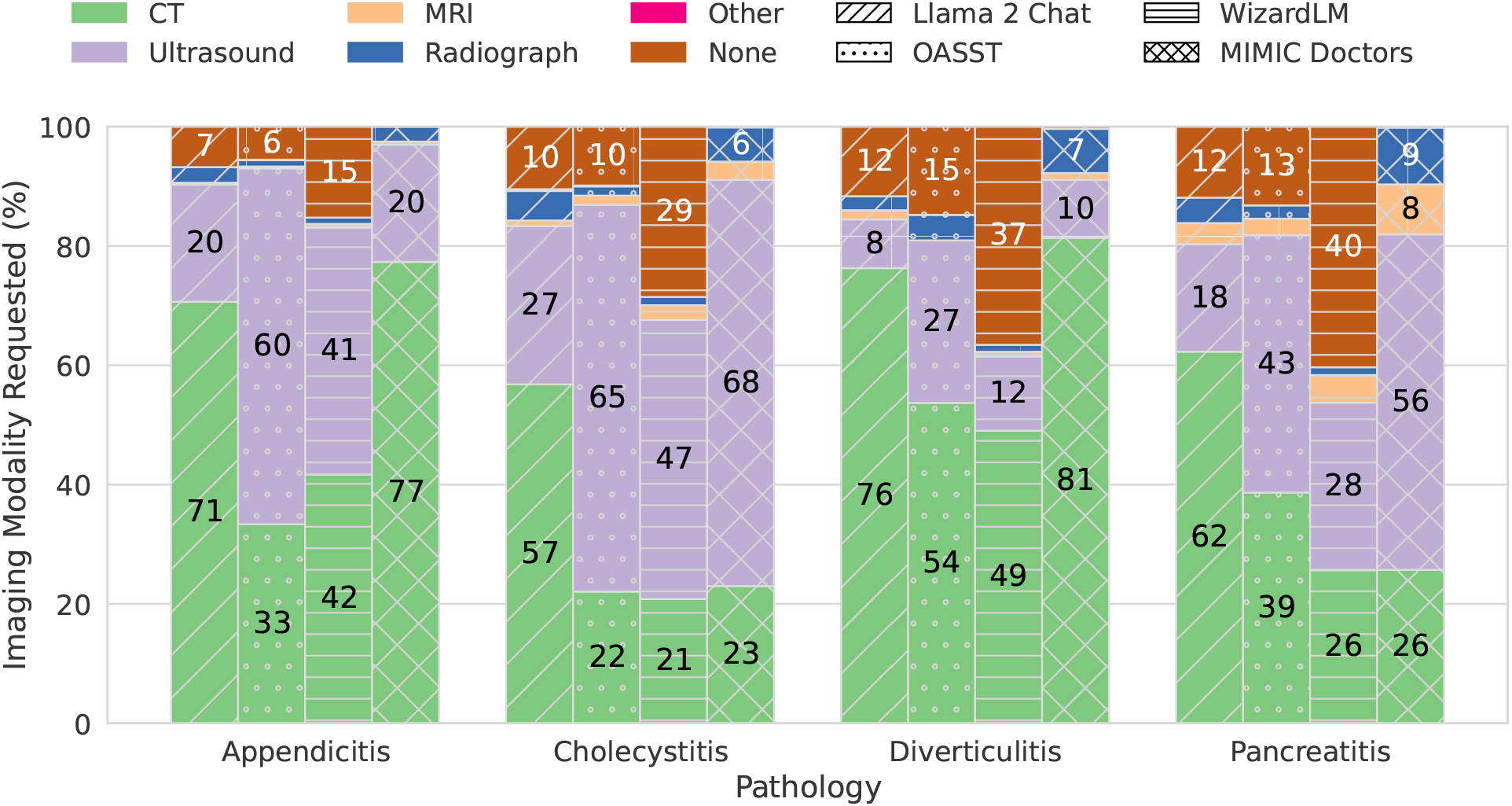
The first imaging modality requested by the LLMs and the attending doctors in the MIMIC dataset are shown. LLMs sometimes follow diagnostic guidelines concerning imaging but often diagnose without requesting any imaging at all. As we show that imaging is the most useful diagnostic tool for all LLMs for each pathology except pancreatitis, this could be partly responsible for their low diagnostic accuracy. The legend specifies the colors of the imaging modalities and the patterns of the models.

## G FI Performance

**Figure 13:**
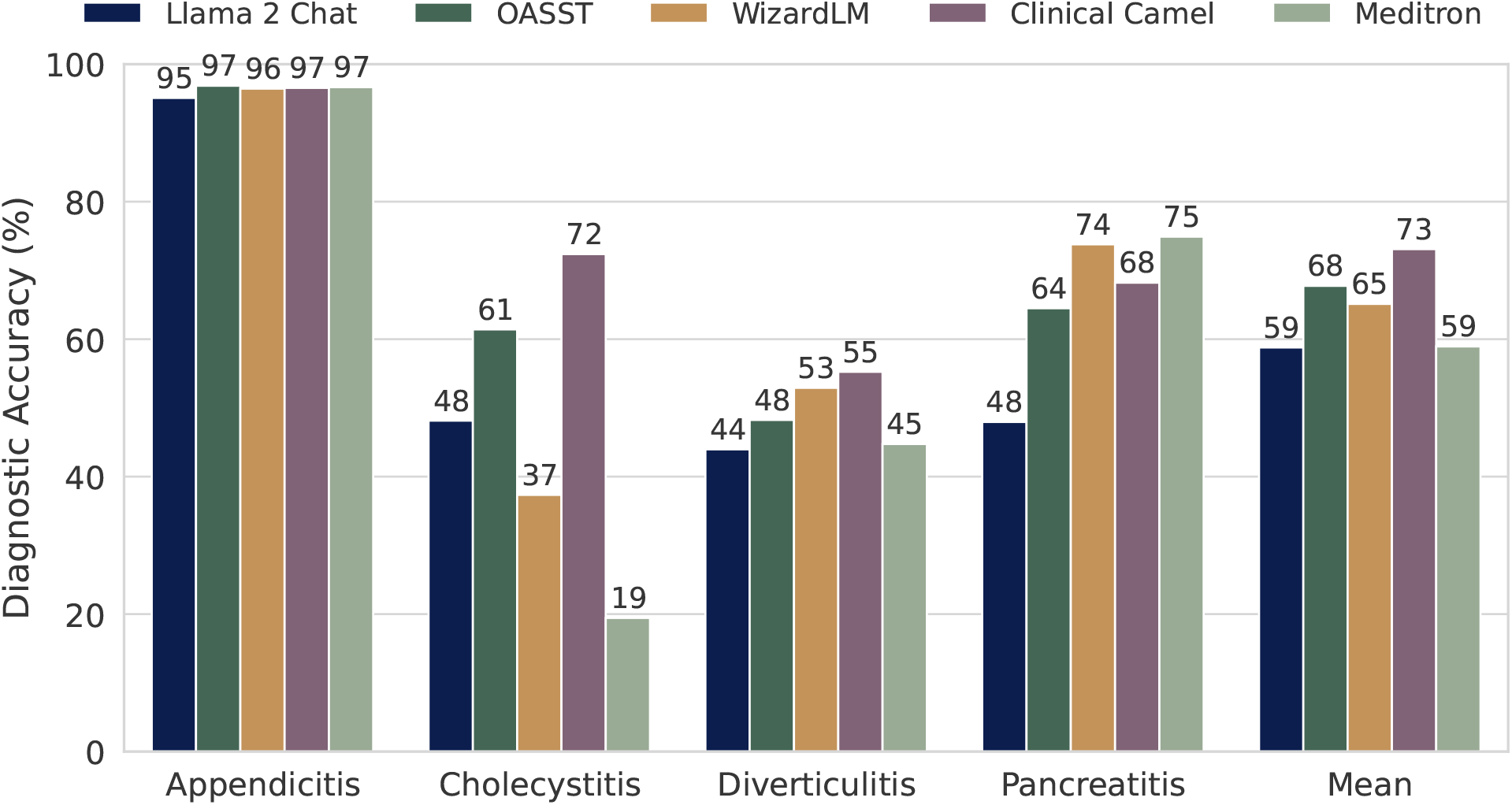
LLMs perform better when all information is given, especially on pathologies with strong indications such as appendicitis (dilated appendix described in radiologist report) and pancreatitis (elevated pancreatic enzymes listed in laboratory test results).

## H LLMs Diagnostic Accuracy Without Medical Abbreviations

**Figure 14:**
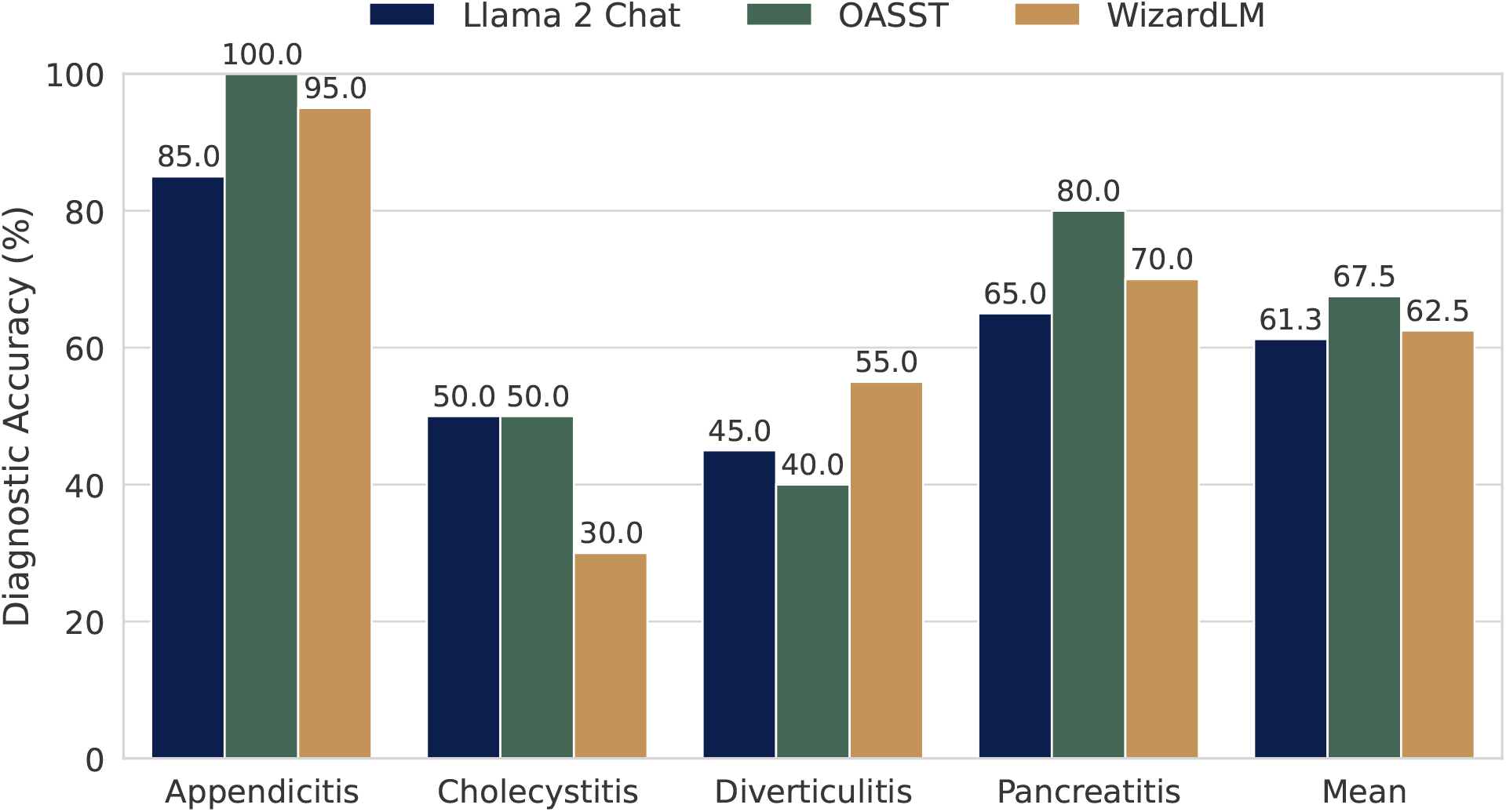
Diagnostic accuracy on the clinician subset of MIMIC-CDM-FI stays the same when medical abbreviations are written out.

## I LLMs are sensitive to information order

Changing the order of the presented information changes diagnostic accuracy. Crucially, the best order is disease specific, meaning a clinician must deliver a preliminary diagnosis to ensure proper model performance, eliminating many of the benefits of an AI clinical decision making system. Best order for each pathology is in bold.

### I.1 Llama 2 Chat

| Experiment<br>Pathology | Imaging, Lab<br>Physical | Imaging, Physical<br>Lab | Lab, Imaging<br>Physical | Lab, Physical<br>Imaging | Physical, Imaging<br>Lab | Physical, Lab<br>Imaging |
|---|---|---|---|---|---|---|
| Appendicitis | **95.72** | **95.72** | 95.61 | 94.88 | 95.40 | 94.98 |
| Cholecystitis | 39.04 | 40.43 | 46.76 | 48.30 | 41.05 | **64.35** |
| Diverticulitis | 45.91 | 46.30 | 41.63 | 42.41 | 47.08 | **49.03** |
| Pancreatitis | 60.41 | **65.99** | 53.16 | 50.37 | 63.01 | 48.14 |
| Mean | 60.27 | 62.11 | 59.29 | 58.99 | 61.64 | **64.13** |

### I.2 OASST

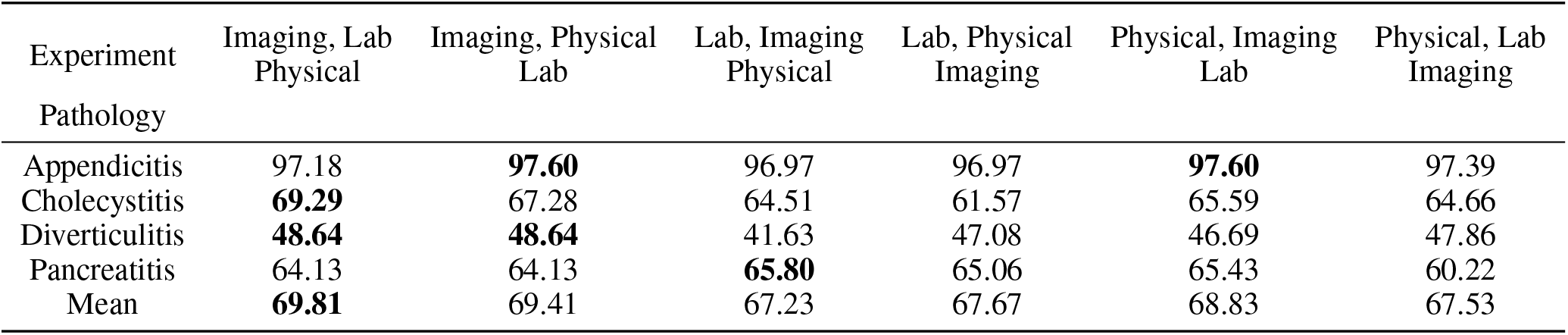

### I.3 WizardLM

| Experiment<br>Pathology | Imaging, Lab Physical | Imaging, Physical Lab | Lab, Imaging Physical | Lab, Physical Imaging | Physical, Imaging Lab | Physical, Lab Imaging |
|---|---|---|---|---|---|---|
| Appendicitis | 96.97 | 96.76 | 96.55 | 96.34 | **97.18** | 96.66 |
| Cholecystitis | 40.74 | 37.19 | 41.82 | 37.96 | 36.42 | **51.39** |
| Diverticulitis | 51.36 | 52.53 | **54.09** | 53.70 | 53.70 | 52.92 |
| Pancreatitis | **78.81** | 77.88 | 75.84 | 75.28 | 78.07 | 69.14 |
| Mean | 66.97 | 66.09 | 67.07 | 65.82 | 66.34 | **67.53** |

## J LLMs Cannot Interpret Numbers

**Figure 15:**
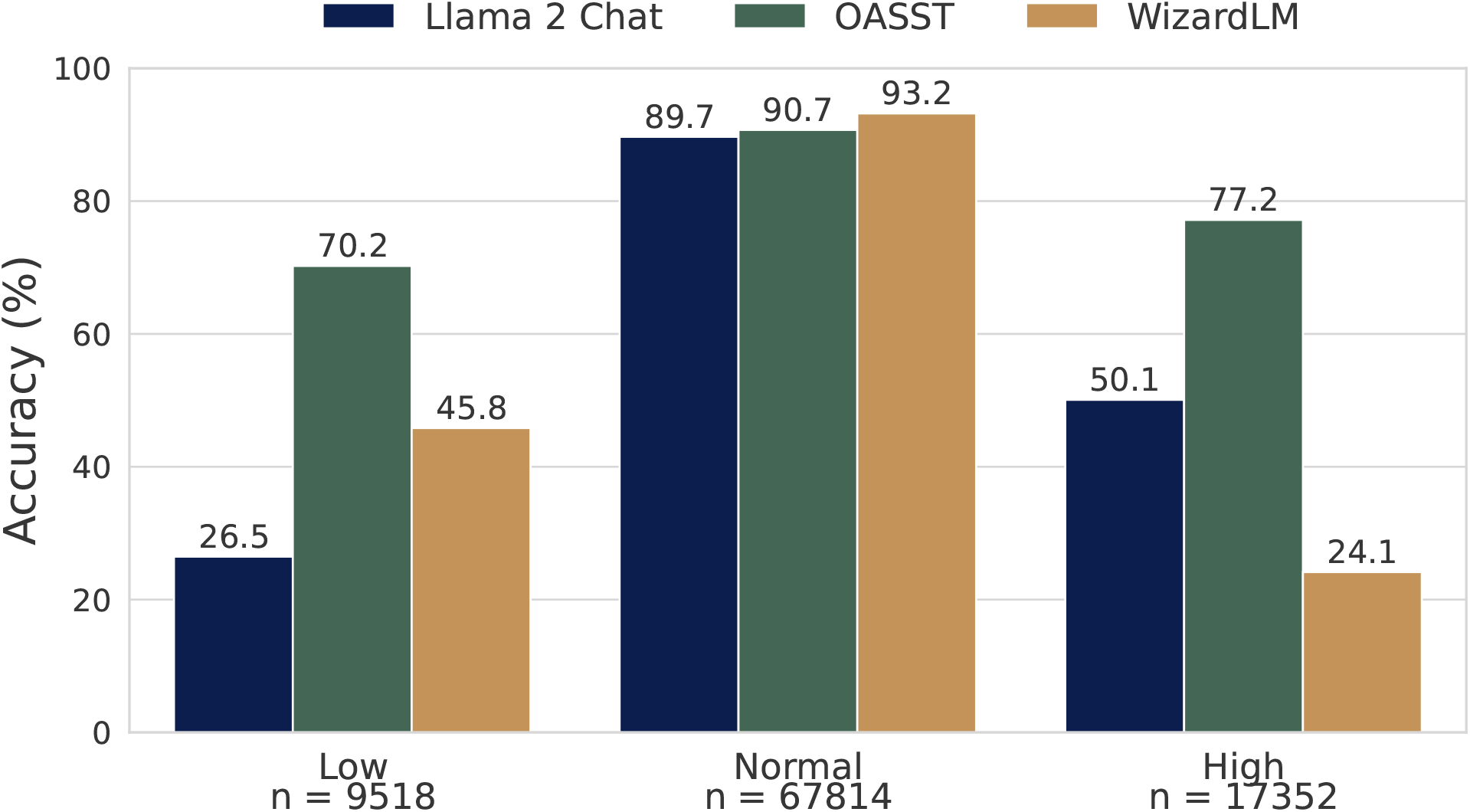
When providing a laboratory test result and its reference range, LLMs are incapable of consistently interpreting the result as normal, low or high.

## K LLMs Struggle to Follow Instructions

**Table 6:**
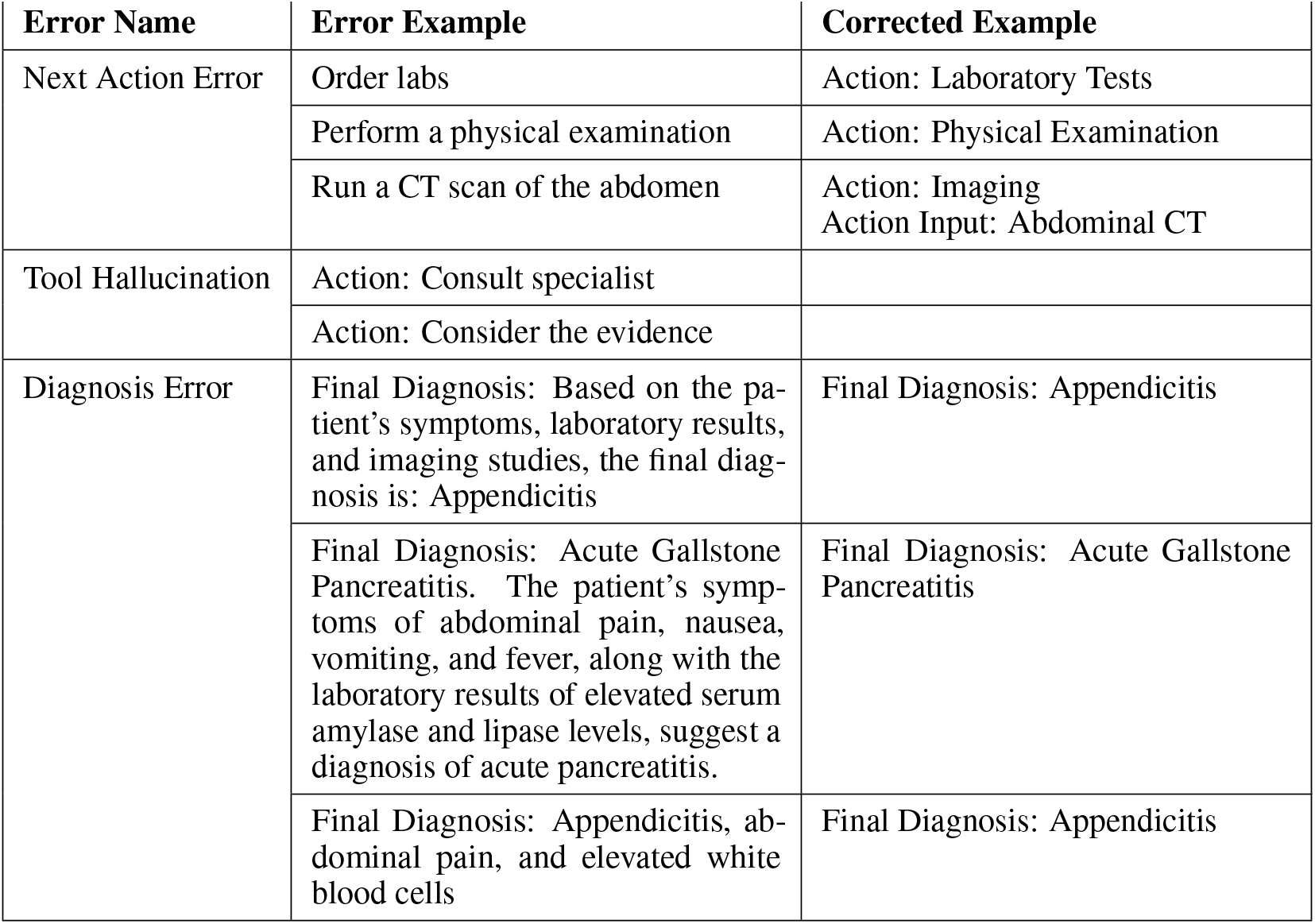
Examples of the types of errors commonly made by models when providing actions and diagnoses. The corrected example in the desired format is also provided. Note that tool hallucination examples are simply not valid actions and so there are no corrected examples provided.

| **Error Name** | **Error Example** | **Corrected Example** |
|---|---|---|
| Next Action Error | Order labs | Action: Laboratory Tests |
|  | Perform a physical examination | Action: Physical Examination |
|  | Run a CT scan of the abdomen | Action: Imaging<br>Action Input: Abdominal CT |
| Tool Hallucination | Action: Consult specialist |  |
|  | Action: Consider the evidence |  |
| Diagnosis Error | Final Diagnosis: Based on the pa-tient’s symptoms, laboratory results, and imaging studies, the final diag-nosis is: Appendicitis | Final Diagnosis: Appendicitis |
|  | Final Diagnosis: Acute Gallstone Pancreatitis. The patient’s symp-toms of abdominal pain, nausea, vomiting, and fever, along with the laboratory results of elevated serum amylase and lipase levels, suggest a diagnosis of acute pancreatitis. | Final Diagnosis: Acute Gallstone Pancreatitis |
|  | Final Diagnosis: Appendicitis, ab-dominal pain, and elevated white blood cells | Final Diagnosis: Appendicitis |

## L LLMs are Sensitive to the Order of Information

**Figure 16:**
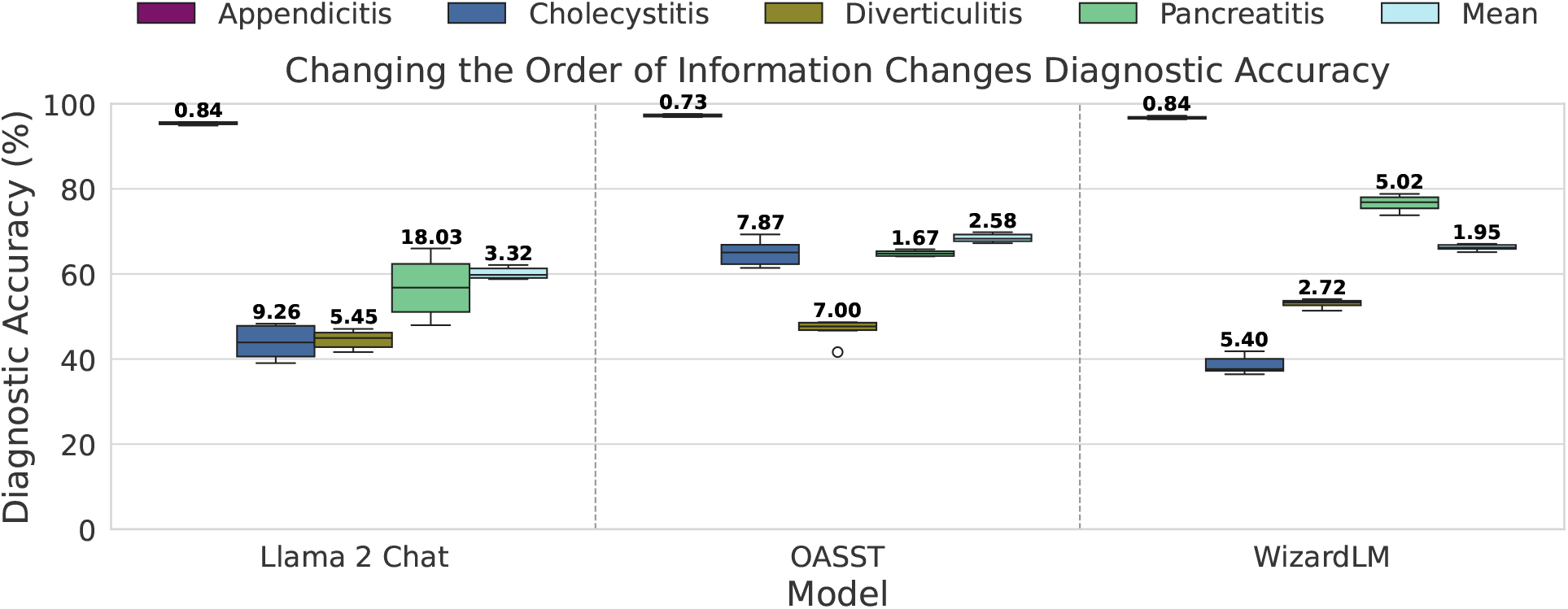
By mixing the order in which information is presented to LLMs, their diagnostic accuracy changes despite the information included staying the same. This places an unnecessary burden upon clinicians who would need to consider and monitor the order in which they feed the models with information. The value above each whisker shows the difference between the best performing and worst performing order.

## M Summarizing Progress Improves CDM Diagnostic Accuracy

**Figure 17:**
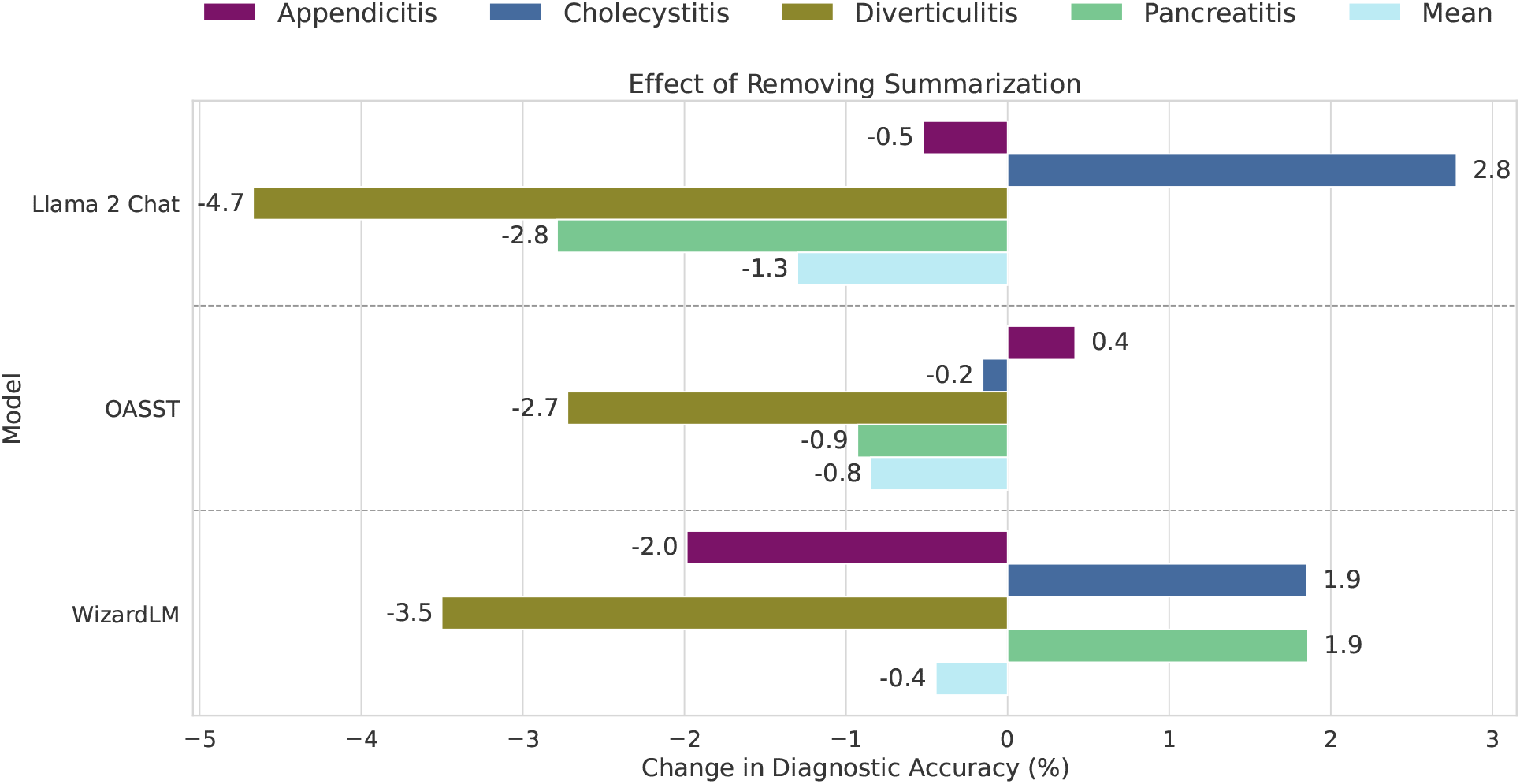
When an LLM approaches its input limit, we ask it to summarize the information gathered thus far to allow for more context. Increased input sizes allows it more opportunities to ask for information and increases the chances of requesting information that is important for the diagnosis.

